# Borderless battles: Modelling the spread of artemisinin partial resistance in connected subpopulations in southern Africa

**DOI:** 10.64898/2026.06.04.26354014

**Authors:** Lovemore Mapahla, Immo Kleinschmidt, Sheetal Prakash Silal

**Affiliations:** The Modelling and Simulation Hub, Africa, Department of Statistical Sciences, University of Cape Town, South Africa; Medical Research Council International Statistics and Epidemiology Group, Department of Infectious Disease Epidemiology, London School of Hygiene and Tropical Medicine, Keppel Street, London WC1E 7HT, United Kingdom; Centre for Global Health, Nuffield Department of Medicine, Oxford University, Oxford, United Kingdom

## Abstract

Artemisinin partial resistance has not yet been reported in southern Africa. Therefore, the magnitude of the spread of artemisinin partial resistance in this region is yet to be quantified.

Using a two strain metapopulation modelling framework, we explored possible spread of artemisinin partial resistance in eight connected countries with high level of human movement. We explored three scenarios in which artemisinin partial resistance may first enter circulation: low malaria transmission level country; high malaria transmission level country and all countries and compared to an artemisinin partial resistance free scenario. Partial rank correlation coefficient sensitivity analysis was performed to identify key parameters that drive artemisinin partial resistance spread.

Our model simulations show that high mobility between countries can increase the spread of mutations associated with delayed clearance. Suggesting that artemisinin partial resistance will be confirmed (>5% partial resistant cases) after 14 years of circulation if it is to appear in southern Africa. We confirm that human movement, both human-to-mosquito and mosquito-to-human probabilities of transmission, were significant and highly sensitive parameters in the spread of artemisinin partial resistance.

Human mobility between countries can facilitate the spread of artemisinin partial resistance. More research is needed to identify strategies to preserve the efficacy of artemisinin-based combination therapies in the presence of partial artemisinin resistance, which may eventually lead to treatment failure and necessitate regimen replacement.

**Author summary:** We developed a two-strain, spatially explicit metapopulation model with eight patches of low, moderate and high levels of malaria transmission. We simulated the spread of artemisinin partial resistance in geographically connected Southern African Development Community Malaria Elimination 8 Initiative (SADC-E8) countries with a high-level of human cross border movement. This study provides a hypothetical reference for future studies necessary to fully understand the spread and impact of artemisinin partial resistance in the malaria elimination process in southern Africa. We found that human movement between countries facilitates the spread of artemisinin partial resistance. The slow parasite clearance and high gametocyte carriage (2-fold of sensitive strain 3 days after ACTs treatment) drive the spread of artemisinin partial resistance within the human population. In the absence of artemisinin partial resistance data in SADC-E8, this study’s simulations inform policy to make decisions on the potential spread of artemisinin partial resistance.

## Introduction

Artemisinin-based combination therapy (ACT) is a combination of an artemisinin derivative and a long-lasting antimalarial compound (partner drug) [1]. ACTs continue to be used as first line treatment for *Plasmodium falciparum* malaria infections across Africa. However, artemisinin partial resistance, which the is delayed clearance of *P. falciparum* parasites following treatment with ACTs, is a threat to progress towards malaria elimination [1–3]. Artemisinin partial resistance is confirmed in a country when a quality-controlled study using ACT finds more than 5% of patients carrying *PfKelch13* resistance-validated mutations and/or experiencing slow clearance and with delayed clearance as shown either by persistent parasitemia detected by microscopy at 72 hours (± 2 hours) after treatment or a parasites clearance half-life ≥5 hours [1]. The delayed parasite clearance increases selective pressure on the partner drug [2], which can drive the emergence of full drug resistance and ultimately lead to full treatment failure, requiring regimen replacement [3].

Artemisinin partial resistance alone rarely leads to treatment failure, but the presence of partner drug resistance brings a high risk of treatment failure. ACTs can still cure patients as long as the partner drug remains effective [2]. In South East Asia, where artemisinin resistant mutations emerged first in early 2000, resistance to several ACTs’ partner drugs were reported [3]. In fact, the loss of first-line ACTs in Western Cambodia were attributed to artemisinin resistant mutations [4]. The main challenge of this situation is the lack of existing and readily available alternative treatment to ACT. Nowadays, there are ongoing trials for alternative antimalarial drugs to ACTs but they may not be available soon and most probably will be more expensive compared to ACTs currently administered free of charge [2, 5]. Therefore, there is need to investigate the potential spread of the yet to be confirmed artemisinin partial resistance in southern African countries who are aiming for malaria elimination [6] to provide a foundation for future research on malaria drug resistance.

Mutations associated with slow clearance can spread geographically through parasite sexual stages in which parasites undergo sexual recombination and maturation, producing offsprings that may carry resistant genes and disperse through transmission during a mosquito bite [7]. In 2025, the World Health Organisation (WHO) reported that artemisinin partial resistance has been confirmed in Eritrea, Rwanda, Uganda and the United Republic of Tanzania, and is suspected in Ethiopia, Namibia, Sudan and Zambia [2]. A de novo emergence of artemisinin resistance mutations was reported in Rwanda [8]. Independent emergence and local spread of artemisinin resistant *P. falciparum* mutations were seen in Uganda, reaching 19.8% prevalence in 2019 [9]. If a *PfKelch13* mutation has only shown to be associated with slow clearance during clinical trial but not validated by in vitro data, it is labelled a candidate marker [10]. More than 20% prevalence of artemisinin resistance candidate markers, the genetic variations that serve as early warnings of slow clearance but yet to be validated [3], was reported in several locations in Uganda in 2022 [11]. The emergence of artemisinin partial resistant parasites can compromise treatment efficacy; potentially reversing progress made toward elimination. It is therefore critical to understand how artemisinin partial resistance spread once it enters southern Africa. Modelling the spread of artemisinin partial resistance in different scenarios of first entrance into circulation can provide insights to support malaria elimination initiatives in southern Africa [12].

In 2009, eight countries in southern Africa formed the Southern African Development Community Malaria Elimination 8 Initiative (SADC-E8) as a collaborative initiative to coordinate malaria control and treatment strategies to achieve elimination by 2030 [6, 13, 14]. The initiative is a coalition of eight countries, Angola, Botswana, Eswatini, Mozambique, Namibia, South Africa, Zambia and Zimbabwe, which are all malaria endemic [2, 13]. Given the geographic connectivity and high population movement between SADC-E8 countries [6], artemisinin-resistant mutations could spread rapidly and potentially undermine the progress made toward malaria elimination.

The dynamics of artemisinin partial resistant malaria transmission can be simulated using mathematical models such as network agent-based models (ABM), stochastic models, pharmacodynamic-pharmacokinetic models, and metapopulation models [15]. Mathematical modelling is a valuable complementary approach to costly field trials, particularly for outcomes that may take a decade or more to be observed [16].

Metapopulation models are spatial models used to investigate the interactions and mobility among different subpopulations across time and space [17]. This type of model was used to assess the dynamics of malaria transmission in Rwanda, Uganda, and the Democratic Republic of Congo (DRC) with human movement [18]. Previous studies have shown that movement of people, including temporary, seasonal, or permanent migration and daily commuting, between areas with different malaria transmission levels, influences the spread of diseases such as malaria [19], and in a metapopulation modelling framework mobility can be included directly or indirectly [20]. Direct (explicit) inclusion of mobility is where the actual movement of individuals between spatial locations is explicitly included in the model whereas indirect (implicit) modelling of movement involves the use of a proxy measure, such as a weighted inverse of distance between populations to mimic actual movement. Given the lack of comprehensive migration data [21], a coupling structure can be used to mimic mobility and indicate the strength of interaction within and between subpopulations. The coupling matrix expresses how subpopulations affect each other’s disease spread.

Several studies have assessed the spread and potential impact of artemisinin partial resistance in Africa. Slater et al. (2016) reported that, across Africa, artemisinin partial and partner drug resistance could result in a 7% increase in malaria cases compared to a scenario without resistance [22]. In Rwanda, an individual-based malaria model was used to evaluate policy interventions to decrease the spread of artemisinin resistance mutations [23]. Scott et al. (2018) predicted that artemisinin partially resistant strains could circulate for up to 10 years longer in areas with high *P. falciparum* prevalence than in low-prevalence areas before resistance is detected in Africa [24]. All these studies are valuable in estimating the spread and impact of artemisinin partial resistance, but not specifically relevant for geographically connected countries in southern Africa. In this contribution, we address this research gap by carrying out simulations for countries that are geographically connected and with high level of cross-border movement [6, 13].

This study aligns with the WHO strategy to respond to antimalarial drug resistance in Africa launched in 2022 [1]. Our scenario simulations have the potential to inform policymakers on strategies to mitigate the spread of artemisinin partial resistance. In particular, our findings may inform efforts within the SADC-E8 region toward strengthening supervision, genomic surveillance, and the enforcement of existing regulations, which could contribute to slowing the spread of artemisinin partial resistance. This study also provides a foundation for future research on malaria drug resistance in connected subpopulations with similar characteristics to those in our study setting.

## Materials and Methods

### Ethics statement

Ethics approval for this study was obtained from Human Research Ethics Committee (HREC) of the University of Cape Town (HREC REF Number: 180/2024).

### Study setting

Our study setting comprises eight countries that are geographically connected with a high level of cross-border movement [6]. *S2 Fig 1* shows the trend of aggregated *P. falciparum* malaria cases in our study setting from 2015 to 2024 and the data was extracted from WHO annexes [2]. The trend plot describes the SADC-E8 region with 2-fold increase in *P. falciparum* malaria cases between 2015 and 2024 with a slight drop in 2021. Malaria transmission levels for our study setting are described in the *S1 Supplementary file*. Artemether lumefantrine (AL) ACT was the primary first-line treatment for *P. falciparum* malaria in all eight countries in 2024 [2]. A full description on the policy to treat malaria with ACTs free of charge in the public sector and free of charge distribution of vector control to all age groups through routine channels is in Table 1 in the *S1 Supplementary file*.

### Model description and formulation

We developed a two-strain, spatially explicit metapopulation model using non-linear ordinary differential equations (ODE) (see Equation (5) and Equation (6) in the *S1 Supplementary file*), to simulate the dynamics of malaria transmission (see *Figure 1*). The population at risk of malaria infection in the SADC-E8 was divided into eight subpopulations: Angola, Botswana, Eswatini, Mozambique, Namibia, South Africa, Zambia, and Zimbabwe. The subpopulations were modelled as patches with malaria state variables described in Table 2, with respective initial conditions in Table 3 in the *S1 Supplementary file.* The initial conditions were derived by running the model with an initial seed for 1 825 days to reach equilibrium conditions. The initial seed of infectious population was 1% of the human population at risk and 1% of mosquito population at risk. The subscripts *r* and *s* are added to both human and mosquito malaria state variables and parameters to represent artemisinin partial resistant and sensitive infections, respectively. Similarly, the subscripts *h* and *v* represent the human and mosquito populations, respectively. The index *i* represents the patch number.

**Figure 1:**
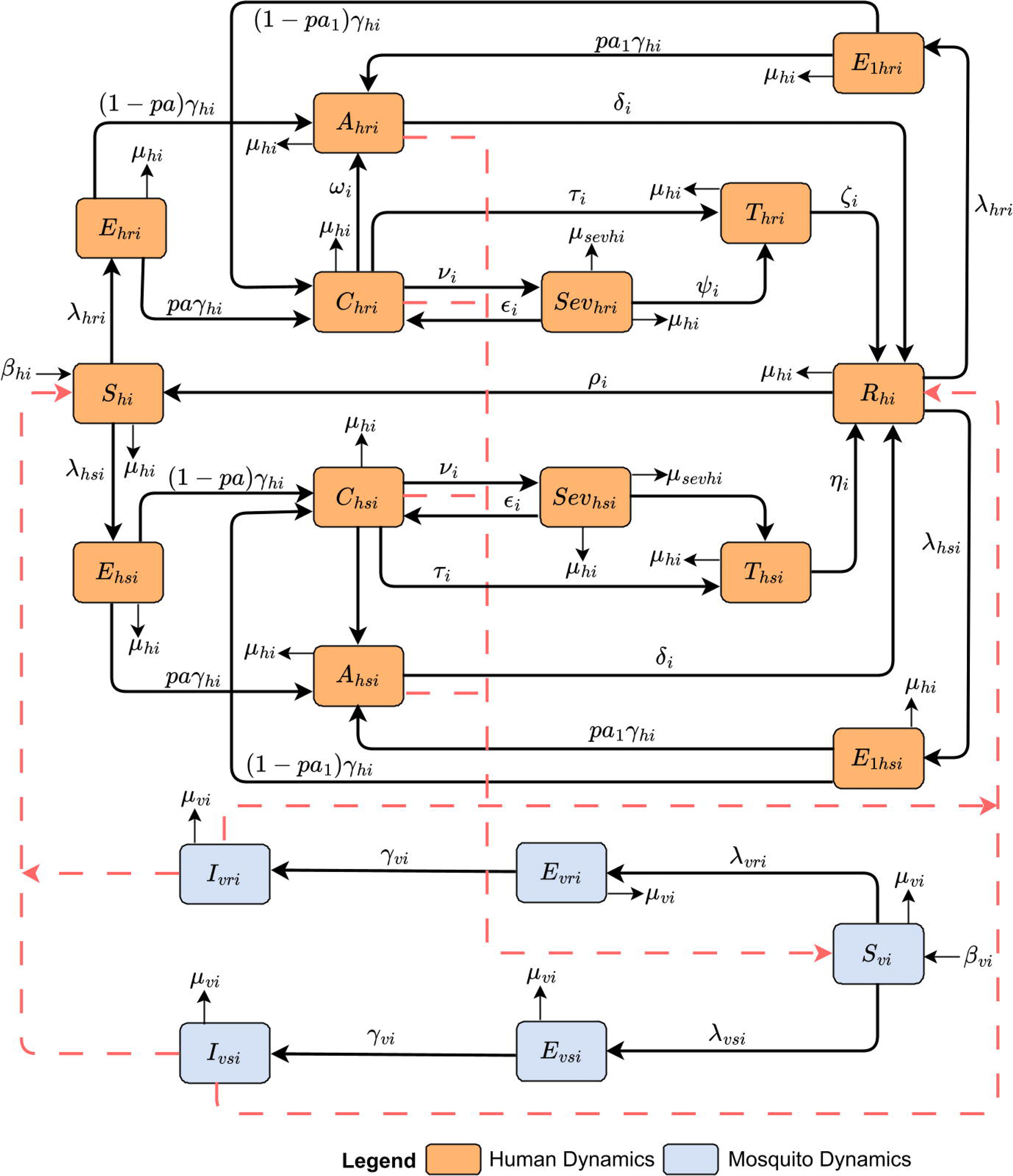
Schematic diagram of malaria transmission between human and mosquito populations in patch i. The dashed red lines show direction of infection after blood meal.

The human total population residing in patch *i*, depending on the disease status, is stratified into 14 epidemiological compartments: the susceptible human population (*S_hi_*), exposed but not infectious (*E_hi_*), asymptomatic but infected (*A_hi_*), symptomatic infected (*C_hi_*), severe cases (*Sev_hi_*), treated (*T_hi_*), recovered (*R_hi_*) and exposed but not infectious (*E*_1*hi*_) after a second blood meal. Except for susceptible and recovered compartments, the compartments are binary with respect to parasite genotype. The probability of having asymptomatic malaria after the second blood meal is *pa*1*_i_* where *pa*1*_i_ > pa_i_*. In the treated human population, parasites will be cleared at a rate of either *η_ri_* or *η_si_*, where *η_ri_ < η_si_*. That is, we assume that artemisinin partial resistant parasites are still cleared by treatment but more slowly.

The adult *Anopheles* mosquito population in each patch is divided into five compartments. Compartments depending on infection status and parasite genotype are the susceptible (*S_Vi_*), exposed (*E_Vi_*) and infectious (*I_Vi_*) mosquito population. Except for the susceptible compartment, the compartments are binary with respect to parasite genotype.

The dynamics of malaria transmission within human and mosquito compartments are governed by the parameters listed in Table 4 in the *S1 Supplementary file*. Our model parameters were chosen based on a review of literature. Human and mosquito population malaria transmission dynamics are linked through the blood meal during mosquito bite.

We used the slow rate of clearance and the gametocyte carriage ratio three days after treatment to drive the spread of artemisinin partial resistant infections. The gametocyte carriage ratio for artemisinin partial resistant to artemisinin sensitive parasites three days after ACTs treatment is approximately 2:1 [25]. We used the Hill equation (with Hill coefficient that controls the steepness, n =1) shown in Equation (7) in the *S1 Supplementary file* and the 3-day gametocyte carriage ratio to estimate the probability of transmission of infection from human to mosquito for the artemisinin partial resistant infections.

Malaria receptivity within patch, determined using the vectorial capacity function, expresses the ability of mosquito species to transmit parasites from infectious human [26–28]. We used Equation (8) in the *S1 Supplementary file* to estimate the vectorial capacity. The vectorial capacity components are assumed to be constant throughout simulation and inferred to parameter estimation in the transmission model.

It was not possible to get temporal human movement data between eight study patches. We used the distance weighted coupling matrix to mimic human movement that is incorporated in two ways: within patch and between patches movements. The latitude and longitude coordinates (shown in Table 5 in the *S1 Supplementary file*) were used to estimate distance between the centroids of patches and create distance matrix – symmetrical distance matrix. Using the formula in Equation (9) and Equation (10) in the *S1 Supplementary file* row weights were obtained. The human interaction between patches is inversely weighted by the distance, so that the closer patches are to each other, the higher the interaction rate is relative to patches that are distant [29]. The denominator was exponentiated to the inverse of movement intensity weight parameter[29], thus either Cl_weight – local mobility intensity or CE_weight – foreign mobility intensity weight. These two parameters determine the degree of linkage within and between patches respectively. Both local and foreign mobility weight intensity are assumed to be constant throughout and inferred to parameter estimation in the transmission model. Sensitivity analysis for the constant movement rate was conducted and presented in *S1 Supplementary file*.

Mosquito survival and biting rate are dependent on seasonal factors. Hence, we incorporated seasonality to reflect the highly seasonal transmission intensity in our study setting [30]. Monthly rainfall data, from 2010 to 2023, obtained from ClimateSERV [31] for each country, was used to determine the respective seasonality function parameters. Equation (11) in the *S1 Supplementary file* was used to derive the parameters. Model assumptions, listed in the *S1 Supplementary file*, were employed to maintain simplicity and interpretability.

### Model data fitting and validation

We split the reported *P. falciparum* malaria cases data extracted from the WHO annexes [2] into two independent data sets: the model fitting (2015–2020) and the model validation (2021-2024) datasets. Vector control coverage data (2000–2024), that is, routine indoor residual spraying (IRS) and insecticide treated nets ITN for each country was extracted from the Malaria Atlas Project (MAP) [32]. The model is run from the year 2000 before being fitted to *P. falciparum* annual reported cases data from 2015 to 2020 using maximum likelihood estimation (MLE) assuming the Poisson distribution with parameter, *λ*, equal to the annual *P. falciparum* reported cases of each country without an offset because total population at specific time is the live time -varying population sum that governs the ODE dynamics at every timestep. Given the unavailability of artemisinin partial resistance cases data SADC-E8, parameter optimisation was done for sensitive cases data only. More details on fitting and validation are described in the *S1 Supplementary file*.

### Model scenarios

We evaluated three scenarios in which artemisinin partial resistant infections may enter circulation: low malaria transmission level setting, high malaria transmission level setting, or multinational and spread. These scenarios were then compared to artemisinin partial resistance free scenario (status quo). We assumed that mosquito control coverage of 2024 remains constant until 2039. A full description of the scenarios is given in Table 7 in the *S1 Supplementary file*. We considered 10%, 50% and 90% where for example 10% migration impact means that 10% of total subpopulation cases are imported cases.

### Sensitivity analysis

We calculated partial rank correlation coefficients (PRCC) using Latin Hypercube Sampling (LHS) [33] to identify the parameters that are important in contributing to the variability in the annual artemisinin partial resistant cases for each country. Uncertainty regarding all parameters bound between 0 and 1 was represented by the beta distribution, while a gamma distribution was used to represent uncertainty regarding all parameters that can take any positive value. A total of 100 computations of each output variable of interest for each country were generated [34]. A tornado plot for PRCC was generated to indicate each parameter’s uncertainty in contributing to the variability in time for the spread of artemisinin partial resistance.

The model code was developed in *R* software [35] and solved using the *deSolve* package. All tests were conducted at a 5% level of significance.

## Results

### Model parameter estimation and model validation

Model parameters and those derived through data fitting are presented in Table 8 in the *S1 Supplementary file*. *S3 Fig 3* shows model validation for the eight SADC-E8 countries.

### Scenario comparisons

Annual malaria incidence (cases per 1,000 people at risk) under different scenarios following the hypothetical introduction of slow-clearance mutations is compared across countries in *Figure 2*. The artemisinin partial resistance free scenario has the least annual incidence case per 1,000 people at risk relative to the other three scenarios. Regardless of the country of first entrance into circulation’s transmission level, artemisinin partial resistance will spread through all connected countries, impacting annual incidence case per 1,000 people at risk. The plots show that there is an earlier and greater impact in low transmission countries such as Botswana, Eswatini, Namibia, and South Africa than high transmission countries such as Angola and Mozambique and Zambia. The daily new infections plots shown in *S4 Fig 4a* - *S4 Fig 4h* show exponentially increasing proportions over 14 years of these delayed clearance phenotype infections in all countries.

**Figure 2:**
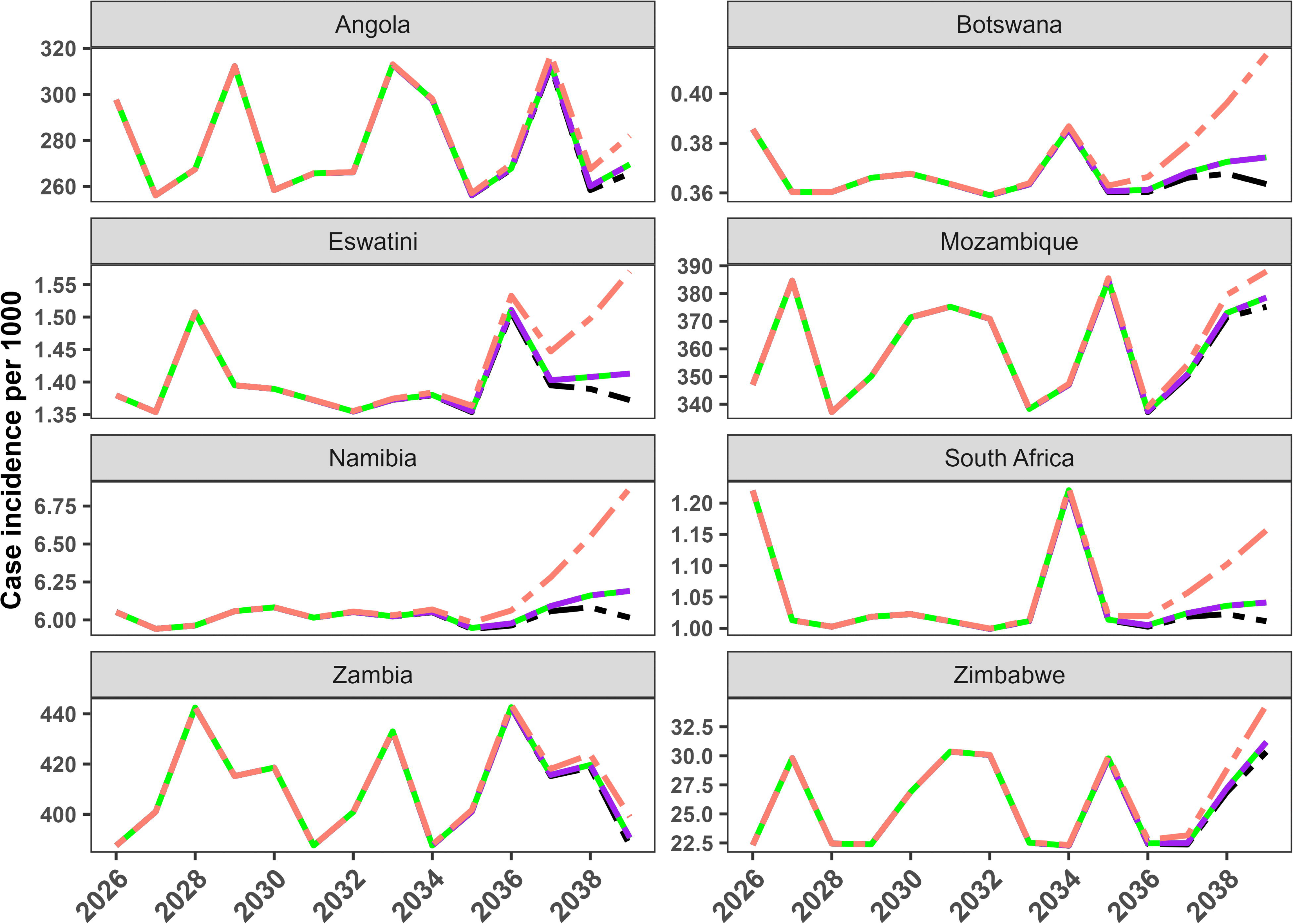
Predicted impact of artemisinin partial resistance on P. falciparum annual case incidence per 1,000 people at risk in eight connected countries, compared with a scenario without artemisinin partial resistance (black dashed line). Scenarios include first hypothetical introduction in low transmission Botswana (green dashed line),, first introduction in high transmission Angola (purple dashed line), and simultaneous multinational introduction (salmon dashed line).

Figure 3 The heatmap showing the progression of artemisinin partial resistance after the first hypothetical introduction in different scenarios *in Figure 3*, reveals that artemisinin partial resistance will be spread and confirmed in all eight connected countries 15 years after introduction, regardless of the country of first introduction. However, the confirmation year differs.When artemisinin partial resistance is first introduced in Botswana, and when artemisinin partial resistance is first introduced in Angola, artemisinin partial resistance will be confirmed (exceed 5% threshold) by 2038 and 2039, respectively. In the worst scenario, where artemisinin resistance is introduced in all countries at the same time, artemisinin partial resistance will be confirmed in all countries by 2036.

**Figure 3:**
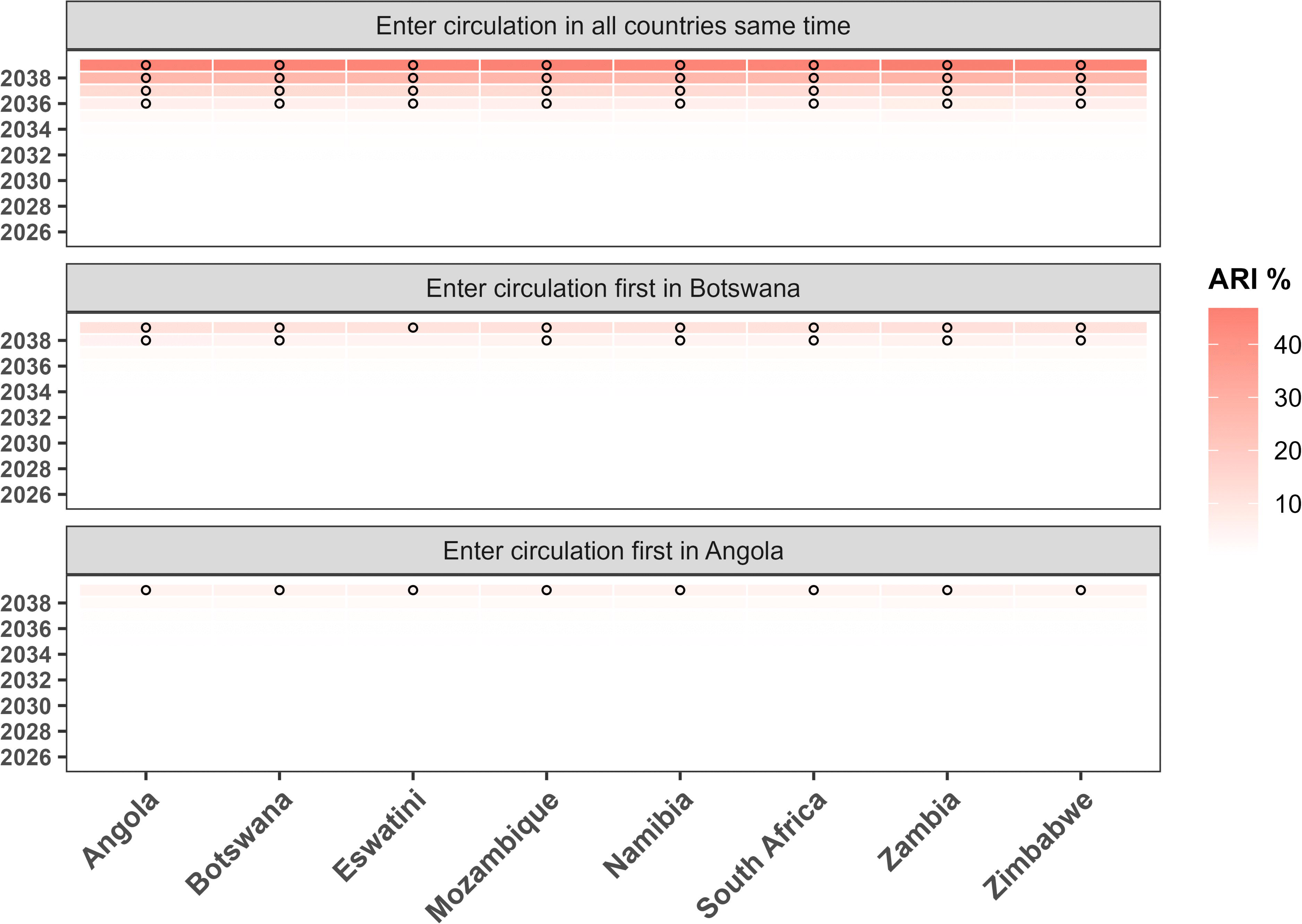
Heatmap for P. falciparum artemisinin partial resistant cases as percentage of total cases since hypothetical introduction. The black circle shows the year in which >5% P. falciparum artemisinin partial resistant cases are reported. APR % is the artemisinin partial resistant cases as a percentage of total cases.

### Assessing different levels of connectivity

The plots in *Figure 4* show that high mobility increases the spread of artemisinin partial resistance. The 10% mobility has the least number of annual artemisinin partial resistance cases compared to the 50% and 90% mobility impact in all countries. Ten years after the hypothetical introduction, by 2034, the three levels of human movement seem to have clear different effects in artemisinin partial resistant cases.

**Figure 4:**
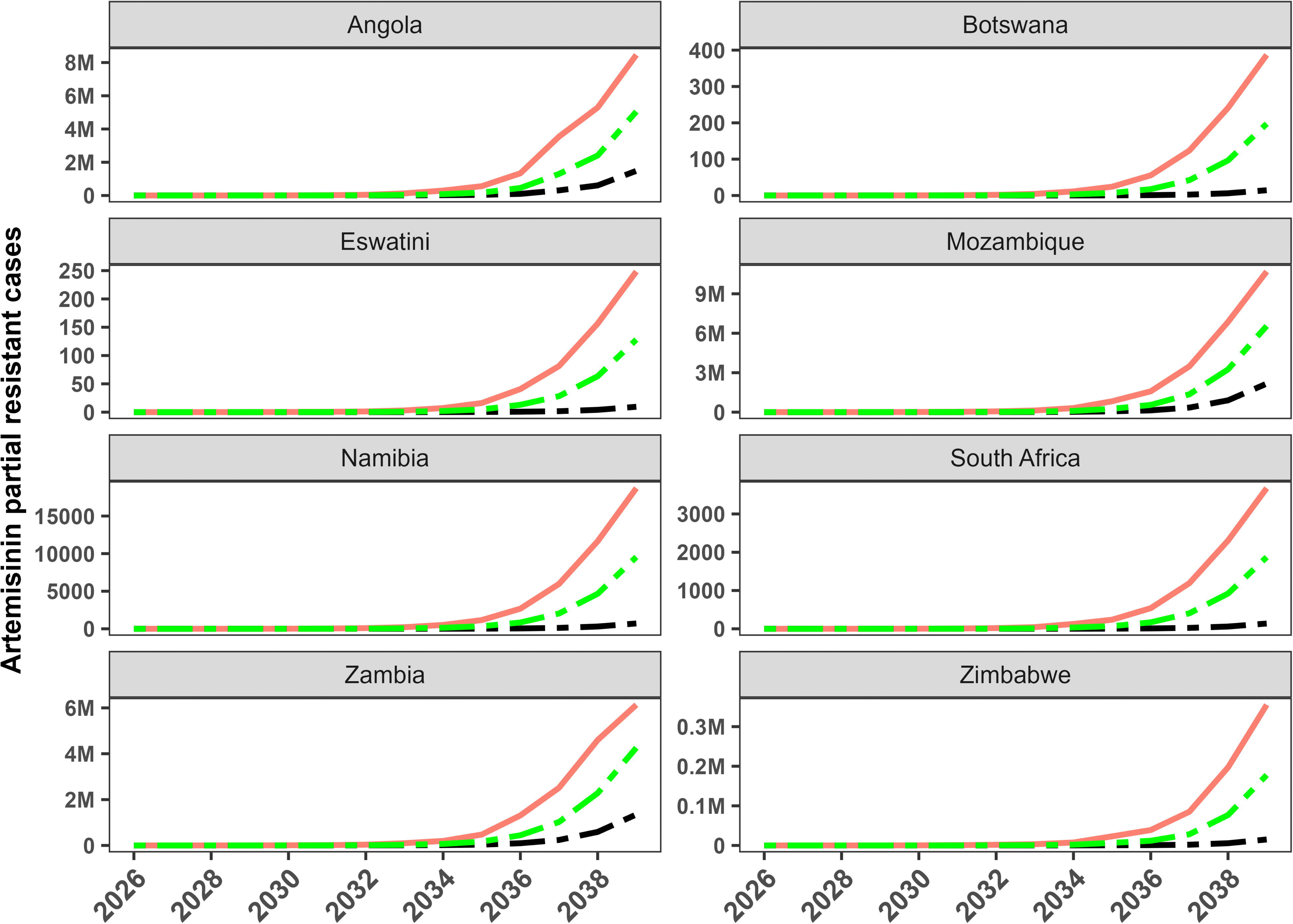
Predicted impact of different mobility levels on the number of P. falciparum artemisinin partial resistant cases in each country. The solid salmon line shows the cases attributed to 90% mobility; the dashed green line shows the cases attributed to 50% mobility impact, while the dashed black line shows the cases attributed to 10% mobility impact.

### Sensitivity analysis

The parameters significantly correlated with the number artemisinin partially resistant cases are shown in Table 12 in the *S1 Supplementary file*. The daily mosquito biting rate and the probabilities of transmission from human to mosquito and mosquito to human, for artemisinin partial resistant infection were significant and highly sensitive parameters in the resistant spread model. The external human mobility weight parameter was positively associated with number of artemisinin partial resistant cases, specifically for low transmission countries. The tornado plots in *S5 Fig 5* show significant parameters (*) and the direction of their associations.

## Discussion

Since 2009, the SADC-E8 has aimed to create a malaria-free region [14, 36]. This aim faces threats that include the potential entrance into circulation of parasites with delayed clearance to ACTs. This slow clearance may increase pressure on partner drugs and ultimately lead to full treatment failure requiring regimen replacement [1, 3].

In this study, we formulated a two-strain, spatially explicit eight-patch malaria compartmental metapopulation model framework. Our simulation results show that artemisinin partial resistance will spread if it enters circulation in at least one of the study subpopulations. When first introduced in Botswana, Angola or all eight countries, simulations show that artemisinin partial resistance would have spread to all countries after 14 years of circulation. In fact, eleven years after the hypothetical introduction in all countries simultaneously, artemisinin partial resistance is predicted to surpass 5% of the total case threshold [1].

Our simulations indicate that human mobility between subpopulations can facilitate the regional spread of artemisinin partial resistance once it emerges in a single country in southern Africa. A high level of human mobility will lead to a greater number of annual artemisinin partial resistance cases relative to a low level of human movement. These findings are similar to those reported in the Greater Mekong Subregion (GMS), where Cambodia, Vietnam, Thailand, Laos, and Myanmar with high level of human population movement are experiencing high proportions of slow clearance [37].

This study highlights the impact of slow clearance on human to mosquito probability transmission. Artemisinin partial resistant gametocyte carriage, that is 2-fold of artemisinin sensitive malaria 72 hours after ACTs treatment [25], increased the probability of transmission for the artemisinin partial resistant strain.

Regardless of which country’s malaria transmission level the artemisinin partial resistance first entered circulation, low transmission countries were characterised by an early and large impact on annual incidence cases per 1,000 compared to high transmission countries. A mathematical modelling study by Scott et al. (2018) had similar findings regarding the relationship between *P. falciparum* prevalence and time-to-emergence and spread of artemisinin resistance [24]. Our findings confirm the need to maintain existing collaborations between SADC-E8 countries to mitigate the impact of artemisinin partial resistance [38].

The study results suggest that over a 14-year timeframe, more than 5% of reported cases in all scenarios and countries would carry mutations associated with delayed clearance. If such mutations were to enter circulation in 2025, artemisinin partial resistance would therefore be confirmed in the study setting by 2039. Scott et al. (2018) used a population epidemiological based mathematical model to examine the potential spread of artemisinin partial resistance in African countries with varying malaria transmission level, grouped by their annual parasitaemia prevalence (APP). The model projected that, had artemisinin partial resistance entered circulation in 2007, it would have taken approximately 13 years to be confirmed across all settings – regardless of transmission intensity. The projection applied to countries with APP below 10% (Botswana, Namibia, South Africa and Zimbabwe), countries (Zambia) with APP between 10% and 25% and countries with APP between 25% and 37% (Angola and Mozambique) [24]. A genomic surveillance study in all 10 Zambian provinces reported varying prevalences of artemisinin partial resistance markers in infections ranging from 5% to 46% [39]. A cross sectional genomic surveillance study in Namibia reported artemisinin partial resistance candidate and validated markers prevalence of 33.2% and 1.2% respectively [40].

Our model was sensitive to the human to mosquito probability of transmission for resistant strain in spreading artemisinin partial resistance. We demonstrated that the slower rate of parasite clearance may result in slightly higher number of malaria cases. A population dynamic mathematical modelling framework by Maude et al. (2009), had similar findings suggesting that parasite slow clearance resulted in more malaria infections [30]. Human feeding rate per mosquito per day is another sensitive parameter in spreading artemisinin partial resistance. A mathematical model by Chitnis et al. (2008) showed that the mosquito biting rate per day is important for initial malaria transmission and a higher biting rate results in a higher number of malaria cases [41]. In our model, the between-countries human movement intensity parameter was also sensitive, in fact, a high level of human movement increases the spread of artemisinin partial resistance. A stochastic, non-linear, ordinary-differential equation model by Silal et al. (2015) showed a similar finding that human movement facilitates the spread of malaria infections and highly threatens early elimination in low transmission settings [29].

We evaluated three possible first entrance into circulation of artemisinin partial resistance in the region: from a low transmission country, a high transmission country and from all eight countries simultaneously. Our findings show that the entrance of artemisinin partial resistance in at least one of the connected countries in the region characterised by a high level of human movement, will lead to the spread of artemisinin partial resistance.

However, our study had limitations. One limitation is that the randomness of the artemisinin partial resistance appearance was not considered. The dynamics of malaria infections were determined by the initial conditions, given that the regional population at risk of malaria was large throughout the simulations, stochasticity would have little impact [42, 43]. The second limitation is that the subpopulation connectivity interaction is limited only to the SADC-E8 countries, while the human mobility network is more complex. We used distance-weighted interaction to mimic migration; however, the pattern of people migration does not rely solely on the state of proximity but also on different pull and push factors like education, business and employment. Lastly, the ITN and routine IRS effectiveness were assumed to be constant, which may not remain valid because insecticide resistance is spreading [44].

## Conclusion

In summary, the results suggest that connectivity between regions or countries can be a vehicle to spread artemisinin resistant infections if it is to enter circulation in at least one of the connected countries. Artemisinin combination treatment still efficient to treat malaria, but the appearance of artemisinin partial resistant infections leads to increased gametocyte carriage and annual malaria incidences within a 15-year time frame. Our scenario analysis in this study could be used to guide long-term decision-making since we compared different scenarios involving the entrance into circulation of artemisinin partial resistance in settings of different levels of transmission and explored the evolutionary process of the artemisinin resistant strain in connected subpopulations.

This study contributes to the SADC-E8’s goal to eliminate malaria. It supports the need to maintain, strengthen and extend collaboration, cooperation, strong government commitment, and transparency across all nations in southern Africa and beyond. Health bodies and international funding institutions need to openly communicate so that the chances of spreading artemisinin partial resistance are mitigated, and early detection is improved. We recommend regional collaboration with robust monitoring and surveillance, efficient testing and treating at cross border points, and extensive use of chemoprevention and vector control. Our recommendations are in line with the SADC-E8, which are the need for dialogue, both within and between subpopulations, for malaria control and elimination [6, 13] and for a multifaceted global coordination strategy to fight drug resistance [2].

## Data Availability

We used publicly available data with links cited in this study. Parameters used in this study were obtained from cited studies and websites.

https://www.who.int/teams/global-malaria-programme/reports/world-malaria-report-2025

## Acknowledgements

This work was wholly supported by the Bill & Melinda Gates Foundation [INV047-048]. Under the grant conditions of the Foundation, a Creative Commons Attribution 4.0 Generic License has already been assigned to the Author Accepted Manuscript version that might arise from this submission. We also acknowledge the support we received from the SADC-E8 [13] countries and their respective National Malaria Control Programs and the Clinton Health Access Initiative (CHAI). The sponsors had no role in study design, data extraction and analysis, writing and editing of the manuscript, and the decision to submit for publication.

## Data availability

We used publicly available data with links cited in this study. Parameters used in this study were obtained from cited studies and websites. The birth and death rate per 1,000 people per year data for each country were obtained from the World Bank Group site [45].

## Author contributions

Lovemore Mapahla (LM), Immo Kleinschmidt (IK) and Sheetal Prakash Silal (SS). LM: Conceptualization, Formal analysis, Investigation, Methodology, Visualization, Writing IK: Conceptualization, Supervision, Validation SS: Conceptualization, Funding acquisition, Resources, Supervision, Validation

## Supporting information

*Appendix 1: S1 Supplementary file*

*Appendix 2: S2 Fig 1*

*Appendix 3: S2 Fig 2a*

*Appendix 4: S2 Fig 2b*

*Appendix 5: S2 Fig 2c*

*Appendix 6: S2 Fig 2d*

*Appendix 7: S2 Fig 2e*

*Appendix 8: S2 Fig 2f*

*Appendix 9: S2 Fig 2g*

*Appendix 10: S2 Fig 2h*

*Appendix 11: S3 Fig 3*

*Appendix 12: S4 Fig 4a*

*Appendix 13: S4 Fig 4b*

*Appendix 14: S4 Fig 4c*

*Appendix 15: S4 Fig 4d*

*Appendix 16: S4 Fig 4e*

*Appendix 17: S4 Fig 4f*

*Appendix 18: S4 Fig 4g*

*Appendix 19: S4 Fig 4h*

*Appendix 20: S5 Fig 5*

