## Supplementary material for "Borderless battles: Modelling the spread of artemisinin partial resistance in connected subpopulations in southern Africa": SI Supplementary file

Lovemore Mapahla*^1^, Immo Kleinschmidt^2^, Sheetal Prakash Silal^1,3^

1. The Modelling and Simulation Hub, Africa, Department of Statistical Sciences, University of Cape Town, South Africa
2. Medical Research Council International Statistics and Epidemiology Group, Department of Infectious Disease Epidemiology, London School of Hygiene and Tropical Medicine, Keppel Street, London WC1E 7HT, United Kingdom
3. Nuffield Department of Medicine, Oxford University, Oxford, United Kingdom

### Materials and methods

#### Study setting

*Figure 1* shows the trend of aggregated *Plasmodium falciparum* malaria reported cases in Southern African Development Community Elimination 8 Initiative (SADC-E8) countries [1] from 2015 to 2024. The trend plot describes our study setting with a 2-fold increase in *P. falciparum* cases since 2015 to 2024 with a slight drop in 2021. The year 2021 was associated with disruptions including reduced mobility, arising from the coronavirus disease 2019 (COVID-19) pandemic, in funding, malaria survailence, treament and vector control distribution [2].

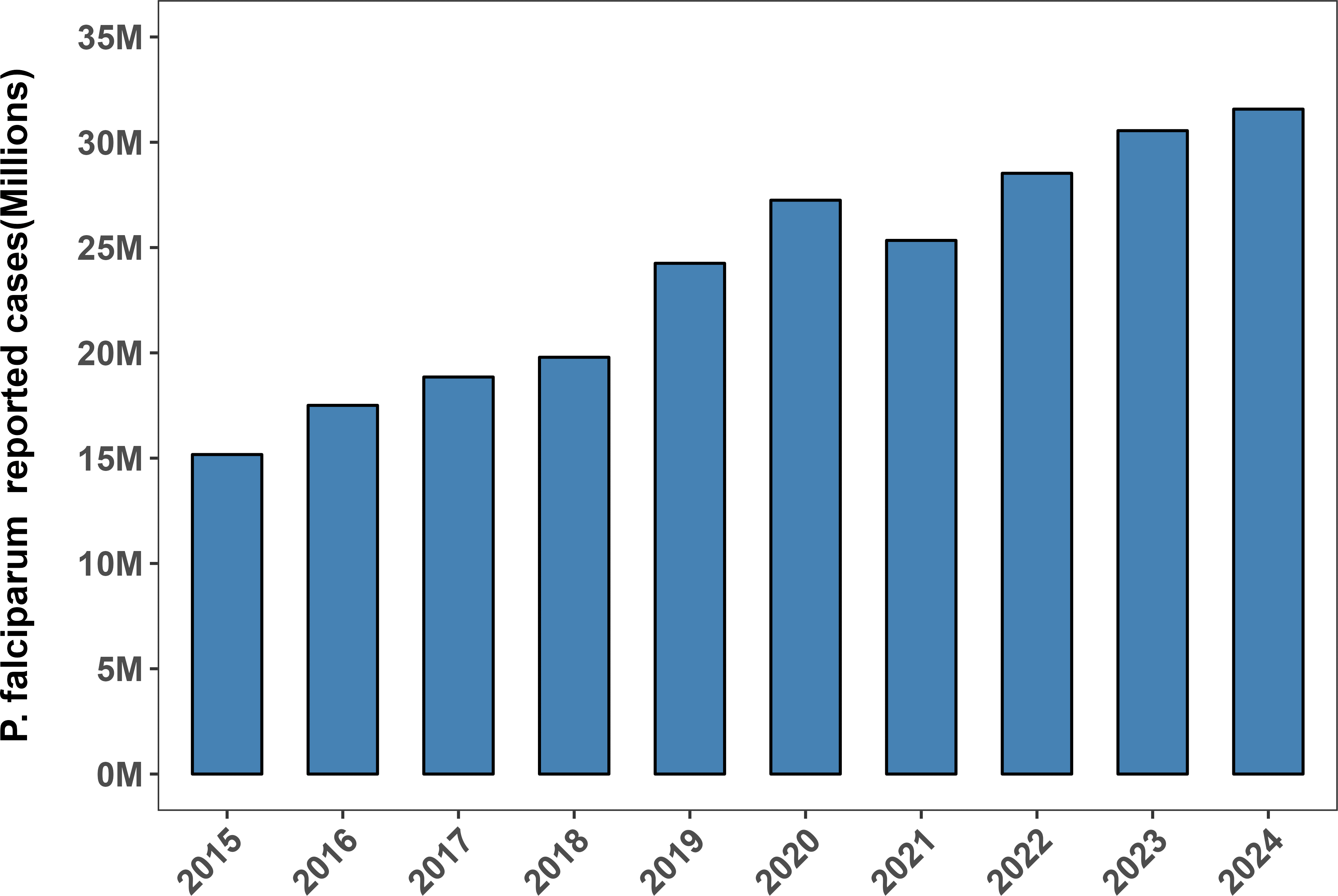

Figure 1: Aggregated P.falciparum malaria cases (2015 to 2024) in millions (M) in SADC-E8 region.

Malaria transmission peaks between March to May in Angola. Angola, which sharesborders with Zambia and Namibia, is characterised by tremendous heterogeneity in malaria prevalence among the 18 provinces [3]. In Botswana, malaria transmission is seasonal and highly unstable and typically peaks between March and April when the country experiences high rainfall and high temperatures [4]. Eswatini that shares borders with South Africa and Mozambique, malaria transmission is seasonal and occurs between November and April of he following year and normally ceases in the cooler months from May to October [5]. Malaria transmission in Mozambique is seasonal throughout the year peaking between December and April of the following year. In Zambia, malaria transmission peaks during the rainy season between November and April, while in Zimbabwe, transmission is seasonal and high between November and June of the following year [6]. The subpopulations experience various malaria transmission levels – Angola (high transmission), Botswana (low transmission), Eswatini (low transmission), Mozambique (high transmission), Namibia (low transmission), South Africa (low transmission), Zambia (high transmission), and Zimbabwe (moderate transmission) [7, 8].

*Table 1* shows transmission levels of our study setting countries based on WHO thresholds. Moderate to high transmission level region is defined as an area with annual parastemia prevalence > 10% or annual parastemia incidence > 250 per 1000 [9]. Botswana , Eswatini, Namibia and South Africa are considered to be frontline malaria elimination countries – low transmission countries[8].

Table 1:Malaria annual incidence case per 1000 in SADC-E8 (2015-2024)

| **Year** | **Angola** | **Botswana** | **Eswatini** | **Mozambique** | **Namibia** | **South Africa** | **Zambia** | **Zimbabwe** |
| --- | --- | --- | --- | --- | --- | --- | --- | --- |
| 2015 | 98.35 | 0.19 | 0.99 | 290.75 | 4.86 | 0.87 | 255.18 | 42.74 |
| 2016 | 130.02 | 0.43 | 0.78 | 311.69 | 10.08 | 0.75 | 286.82 | 27.54 |
| 2017 | 128.16 | 1.22 | 1.36 | 316.73 | 27.27 | 4.06 | 315.67 | 40.38 |
| 2018 | 164.57 | 0.35 | 2.10 | 320.24 | 14.94 | 1.63 | 280.39 | 22.49 |
| 2019 | 217.91 | 0.11 | 0.77 | 392.68 | 1.11 | 0.81 | 278.03 | 25.63 |
| 2020 | 219.54 | 0.56 | 0.70 | 367.68 | 5.68 | 0.74 | 426.10 | 36.59 |
| 2021 | 241.10 | 0.44 | 1.49 | 318.40 | 5.82 | 0.26 | 345.30 | 10.70 |
| 2022 | 220.54 | 0.26 | 0.63 | 379.33 | 4.49 | 0.33 | 403.26 | 11.15 |
| 2023 | 247.57 | 0.34 | 1.73 | 382.64 | 5.01 | 0.84 | 401.40 | 19.13 |
| 2024 | 280.80 | 0.12 | 0.50 | 335.36 | 1.30 | 0.09 | 434.45 | 4.28 |

#### Study setting, mosquito control and malaria treatment policies

The policy to treat *P. falciparum* malaria with ACTs free of charge in 2024 was available and implemented in all countries as indicated *Table 2* [10]**.** All eight countries administered artemether lumefantrine (AL) as the first line treatment plus either primaquine (PQ) or artemether (AM) for *P. falciparum* malaria [10]. In 2024, the policy of free of charge distribution of ITNs to at risk populations was available in Angola, Botswana, Mozambique, Zambia and Zimbabwe. However, the policy was not implemented in Botswana and Zimbabwe [10]. The policy for routine IRS was available free of charge in targeted areas in all countries except in Angola in 2024 [10].

Table 2: Mosquito control and ACTs treatment policy

| **Country** | **Free ITN** | **Free IRS** | **Free ACTs** | **ACTs available** |
| --- | --- | --- | --- | --- |
| **Angola** | Yes | Yes | Yes | AL, DHA-PPQ, AS+AQ, AS |
| **Botswana** | Yes | Yes | Yes | AL-PQ, AS |
| **Eswatini** | No | Yes | Yes | AL, AS |
| **Mozambique** | Yes | Yes | Yes | AL, AS+AQ, AS |
| **Namibia** | No | Yes | Yes | AL+PQ, AS |
| **South Africa** | No | Yes | Yes | AL, AL-PQ, AS, QN |
| **Zambia** | Yes | Yes | Yes | AL, AS |
| **Zimbabwe** | Yes | Yes | Yes | AL, AS |

***Abbreviations:*** *AL: artemether-lumefantrine; AL-PQ: artemether-lumefantrine + primaquine; AM: artemether; AS: artesunate; AS+AQ: artesunate + amodiaquine; DHA-PPQ: dihydroartemisinin-piperaquine; QN: quinine. Co-blistered products are denoted by “+” and co-formulated products are denoted by “-“ [10] .*

### Model equations

#### Human and mosquito population malaria dynamics equations

The human population is divided into 14 compartments while mosquito population is divided into five compartments depending on disease status and the compartments are fully described in *Table 3*. The total human population and total mosquito population at risk of malaria infection in each country at a time point $t$ is shown in *Equation (1)* and *Equation (2)*.

$$\begin{matrix} N_{hi}\left( t \right) & =S_{hi}\left( t \right)+E_{hri}\left( t \right)+E_{hsi}\left( t \right)+A_{hri}\left( t \right)+A_{hsi}\left( t \right)+C_{hri}\left( t \right)+C_{hsi}\left( t \right) \\ & + Sev_{hri}\left( t \right)+Sev_{hsi}\left( t \right)+T_{hri}\left( t \right)+T_{hsi}\left( t \right)+R_{hi}\left( t \right)+E_{1hri}\left( t \right)+E_{1hsi}\left( t \right) \end{matrix}$$

(1)

$$\begin{matrix} N_{vi}\left( t \right) & =S_{vi}\left( t \right)+E_{vri}\left( t \right)+E_{vsi}\left( t \right)+I_{vri}\left( t \right)+I_{vsi}\left( t \right) \\ & \end{matrix}$$

(2)

The non-linear ordinary differential equations in *Equation (5)* and *Equation (6)* were used to simulate the dynamics of malaria transmission in human and mosquito population respectively.

The forces of infection for human popultion, $\lambda_{hri}$ and $\lambda_{hsi}$, given in *Equation* (3)*,* are functions of the levels of vector control, vectorial capacity, probability of transmission from mosquito to human, the proportion of infectiousness in the population and connectivity effect. The forces of infection for mosquito population, $\lambda_{vri}$ and $\lambda_{vsi}$, shown in *Equation (4)* for each patch $i$ are functions of the levels of mosquito control, vectorial capacity, probability of transmission from human to mosquito, the proportion of infectiousness in the population and connectivity effect.

$$\begin{matrix} \lambda_{hri}=\frac{seas*vc*b*connectivity*\left( I_{vri} \right)}{N_{hi}} \\ \lambda_{hsi}=\frac{seas*vc*b*connectivity*\left( I_{vsi} \right)}{N_{hi}} \end{matrix}$$

(3)

$$\begin{matrix} \lambda_{vri} & =\frac{seas*a*c*connectivity*\left( \zeta_{ar}*A_{hri}+C_{hri}+Sev_{hri}+\zeta_{tr}*T_{hri} \right)}{N_{hi}} \\ \lambda_{vsi} & =\frac{seas*a*c*connectivity*\left( \zeta_{as}*A_{hsi}+C_{hsi}+Sev_{hsi}+\zeta_{ts}*T_{hsi} \right)}{N_{hi}} \end{matrix}$$

(4)

$$\begin{matrix} \frac{dS_{hi}}{dt} & =\beta_{hi}N_{hi}\left( t \right)-\lambda_{hri}S_{hi}\left( t \right)-\lambda_{hsi}S_{hi}\left( t \right)-\mu_{hi}S_{hi}\left( t \right)+\rho R_{hi}\left( t \right) \\ \frac{dE_{hri}}{dt} & =\lambda_{hri}S_{hi}\left( t \right)-pa\gamma_{h}E_{hri}\left( t \right)-\left( 1-pa \right)\gamma_{h}E_{hri}\left( t \right)-\mu_{hi}E_{hri}\left( t \right) \\ \frac{dE_{hsi}}{dt} & =\lambda_{hsi}S_{hi}\left( t \right)-pa\gamma_{h}E_{hsi}\left( t \right)-\left( 1-pa \right)\gamma_{h}E_{hsi}\left( t \right)-\mu_{hi}E_{hsi}\left( t \right) \\ \frac{dA_{hri}}{dt} & =pa\gamma_{hi}E_{hri}\left( t \right)+\omega C_{hri}\left( t \right)-\mu_{hi}A_{hri}\left( t \right)-\delta A_{hri}\left( t \right)+pa_{1}\gamma_{h}E_{1hri}\left( t \right) \\ \frac{dA_{hsi}}{dt} & =pa\gamma_{h}E_{hsi}\left( t \right)+\omega C_{hsi}\left( t \right)-\mu_{hi}A_{hsi}\left( t \right)-\delta A_{hsi}\left( t \right)+pa_{1}\gamma_{h}E_{1hsi}\left( t \right) \\ \frac{dC_{hri}}{dt} & =\left( 1-pa \right)\gamma_{h}E_{hri}\left( t \right)+\epsilon Sev_{hri}\left( t \right)-\omega C_{hri}\left( t \right)-\mu_{hi}C_{hri}\left( t \right)-\tau C_{hri}\left( t \right) \\ & -\nu C_{hri}\left( t \right)+\left( 1-pa_{1} \right)\gamma_{h}E_{1hri}\left( t \right) \\ \frac{dC_{hsi}}{dt} & =\left( 1-pa \right)\gamma_{h}E_{hsi}\left( t \right)+\epsilon Sev_{hsi}\left( t \right)-\omega C_{hsi}\left( t \right)-\mu_{hi}C_{hsi}\left( t \right)-\tau C_{hsi}\left( t \right) \\ & -\nu C_{hsi}\left( t \right)+\left( 1-pa_{1} \right)\gamma_{h}E_{1hsi}\left( t \right) \\ \frac{dSev_{hri}}{dt} & =\nu C_{hri}\left( t \right)-\epsilon Sev_{hri}\left( t \right)-\psi Sev_{hri}\left( t \right)-\mu_{sevh}Sev_{hri}\left( t \right)-\mu_{hi}Sev_{hri}\left( t \right) \\ \frac{dSev_{hsi}}{dt} & =\nu C_{hsi}\left( t \right)-\epsilon Sev_{hsi}\left( t \right)-\psi Sev_{hsi}\left( t \right)-\mu_{sevh}Sev_{hsi}\left( t \right)-\mu_{hi}Sev_{hsi}\left( t \right) \\ \frac{dT_{hri}}{dt} & =\tau C_{hri}\left( t \right)+\psi Sev_{hri}\left( t \right)-\mu_{hi}T_{hri}\left( t \right)-\eta_{r}T_{hri}\left( t \right) \\ \frac{dT_{hsi}}{dt} & =\tau C_{hsi}\left( t \right)+\psi Sev_{hsi}\left( t \right)-\mu_{hi}T_{hsi}\left( t \right)-\eta_{s}T_{hsi}\left( t \right) \\ \frac{dR_{hi}}{dt} & =\eta_{s}T_{hsi}\left( t \right)+\eta_{r}T_{hri}\left( t \right)+\delta A_{hri}\left( t \right)+\delta A_{hsi}\left( t \right)-\mu_{hi}R_{hi}\left( t \right)-\rho R_{hi}\left( t \right) \\ & -\lambda_{hsi}R_{hi}\left( t \right)-\lambda_{hri}R_{hi}\left( t \right) \\ \frac{dE_{1hri}}{dt} & =\lambda_{hri}R_{hi}\left( t \right)-pa_{1}\gamma_{h}E_{1hri}\left( t \right)-\left( 1-pa_{1} \right)\gamma_{h}E_{1hri}\left( t \right)-\mu_{hi}E_{1hri}\left( t \right) \\ \frac{dE_{1hsi}}{dt} & =\lambda_{hsi}R_{hi}\left( t \right)-pa_{1}\gamma_{h}E_{1hsi}\left( t \right)-\left( 1-pa_{1} \right)\gamma_{h}E_{1hsi}\left( t \right)-\mu_{hi}E_{1hsi}\left( t \right) \end{matrix}$$

(5)

$$\begin{matrix} \frac{dS_{vi}}{dt} & =\beta_{v}N_{vi}\left( t \right)-\left( \mu_{vi}+\lambda_{vri}+\lambda_{vsi} \right)S_{vi}\left( t \right) \\ \frac{dE_{vri}}{dt} & =\lambda_{vri}S_{vi}\left( t \right)-\left( \mu_{vi}+\gamma_{vi} \right)E_{vri}\left( t \right) \\ \frac{dE_{vsi}}{dt} & =\lambda_{vsi}S_{vi}\left( t \right)-\left( \mu_{vi}+\gamma_{vi} \right)E_{vsi}\left( t \right) \\ \frac{dI_{vri}}{dt} & =\gamma_{vi}E_{vri}\left( t \right)-\mu_{vi}I_{vri}\left( t \right) \\ \frac{dI_{vsi}}{dt} & =\gamma_{vi}E_{vsi}\left( t \right)-\mu_{vi}I_{vsi}\left( t \right) \end{matrix}$$

(6)

Transmission probabilities from treated humans with artemisinin partial resistant infections are higher than those from treated humans with sensitive infections [11] . The slow clearance after ACTs treatment [12] and increased gametocyte carriage for the artemisinin partial resistant strain [13] also drive the spread of artemisinin partial resistance. Malaria infection rises with increasing gametocyte carriage, well described by the Gompertz model reported by Churcher et al. (2013) [14]. Gametocytaemia level in human blood is highly correlated to the probability of human-to-mosquito transmission [15]. If infected people are untreated, artemisinin-sensitive parasites will outcompete artemisinin partial resistant parasites. However, in ACTs-treated hosts, artemisinin partial resistant parasites have selective advantage [16]. After ACTs treatment, gametocyte carriage is 2-fold among patients with slow parasite clearance relative to sensitive parasites suggesting higher probability of transmission [13].

Human-to-mosquito probability of transmission is not directly proportional to gametocyte carriage [14] hence doubling probability of transmission for sensitive strain to get probability of transmission for artemisinin partial resistant strain is unrealistic – it ignores saturation. The relationship between gametocyte carriage and mosquito infection probability is nonlinear (hyperbolic) and best described using the Hill equation [15, 17, 18]. The Hill equation permit the estimation of the probability of an adverse outcome for a given dosage of therapy [17] and modelers are using it to determine the probability of transmission of infection [15].

In this study we used the Hill equation (with hill coefficient that controls the steepness, n =1) shown in *Equation (7)* to estimate the probability of transmission of infection from human to mosquito for the artemisinin partial resistant infections conditional on the probability of transmission of infection from human to mosquito for the sensitive infections and the 3-day gametocyte ratio [13]. This sigmoidal function ensures that the probability of transmission of infection from human to mosquito for the artemisinin partial resistant infections remains between 0 and 1 [19].

$$\begin{matrix} P_{r}=\frac{G^{n}}{G^{n}+\left( G_{50} \right)^{n}} \end{matrix}$$

(7)

where $P_{r}$ is the malaria transmission probability during a blood meal, *G* is the gametocyte carriage in the human host blood 3 days after ACTs treatment, n is the Hill coefficient (n=1) that determines the steepness of the sigmoidal curve and $G_{50}$ is the gametocyte carriage for which $P_{r}$=50%, it is the half-maximal infectious gametocyte carriage representing the specific gametocyte carriage at which 50% of the maximum probability is achieved.

We used receptivity to estimate variations in malaria risk where $VC_{i}$ is the vectorial capacity in *Equation (8)*, $m_{i}$ is the relative density of mosquitoes in relation to human density, $p_{i}$ is the proportion of mosquitoes surviving per day in patch $i$ and $n_{vi}$ represents the parasite’s extrinsic incubation period or extrinsic cycle in days [20].
$p_{i}$ is the exponent of negative mosquito death mortality rate per day (-
$\mu_{vi}$).

$$\begin{matrix} VC_{i}=\frac{m_{i}a^{2}{p_{i}}^{n_{vi}}}{-ln\left( p_{i} \right)} \end{matrix}$$

(8)

The model state variables and initial conditions are described in detail in *Table 3* and *Table 4* respectively while model parameters are in *Table 5.*

Table 3: Description of the state variables of model equation systems for both human and mosquito populations

| **Variable** | **Description** |
| --- | --- |
| $N_{hi}$ | Human population at risk to malaria infection in patch $i$ |
| $S_{hi}$ | Susceptible or malaria naive human population in patch $i$ |
| $E_{hri}$ | Artemisinin resistant strain infected but not infectious human population in patch $i$ |
| $E_{hsi}$ | Artemisinin sensitive strain infected but not infectious human population in patch $i$ |
| $A_{hri}$ | Artemisinin resistant strain infected and asymptomatic human population in patch $i$ |
| $A_{hsi}$ | Artemisinin sensitive strain infected and asymptomatic human population in patch $i$ |
| $C_{hri}$ | Artemisinin resistant strain infected and symptomatic human population patch $i$ |
| $C_{hsi}$ | Artemisinin sensitive strain infected and symptomatic human population patch $i$ |
| ${Sev}_{hri}$ | Artemisinin resistant strain infected human population with severe malaria patch $i$ |
| ${Sev}_{hsi}$ | Artemisinin sensitive strain infected human population with severe malaria patch $i$ |
| $T_{hri}$ | Human population with artemisinin resistant strain who sought treatment in patch $i$ |
| $T_{hsi}$ | Human population with artemisinin sensitive strain who sought treatment in patch $i$ |
| $R_{i}$ | Recovered human population in patch $i$ |
| $E_{1hri}$ | Artemisinin resistant strain infected but not infectious human population in patch $i$ after second blood meal |
| $E_{1hsi}$ | Artemisinin sensitive strain infected but not infectious human population in patch $i$ after second blood meal |
| $N_{vi}$ | Mosquito population in patch $i$ |
| $S_{vi}$ | Susceptible mosquito population in patch $i$ |
| $E_{vri}$ | Artemisinin resistant strain infected but not infectious mosquito population in patch $i$ |
| $E_{vsi}$ | Artemisinin sensitive strain infected but not infectious mosquito population in patch $i$ |
| $I_{vri}$ | Artemisinin resistant strain infectious mosquito population in patch $i$ |
| $I_{vsi}$ | Artemisinin sensitive strain infectious mosquito population in patch $i$ |

Table 4: Initial states values

| **State** | **Angola** | **Botswana** | **Eswatini** | **Mozambique** | **Namibia** | **South Africa** | **Zambia** | **Zimabwe** |
| --- | --- | --- | --- | --- | --- | --- | --- | --- |
| $S_{hi}$ | 16 390 062 | 1 144 556 | 288 138 | 17 764 504 | 1 443 543 | 4 680 926 | 9 887 136 | 9 314 943 |
| $E_{hri}$ | 0 | 0 | 0 | 0 | 0 | 0 | 0 | 0 |
| $E_{hsi}$ | 40 000 | 2 800 | 720 | 44 411 | 3 608 | 11 702 | 24 717 | 23 287 |
| $A_{hri}$ | 0 | 0 | 0 | 0 | 0 | 0 | 0 | 0 |
| $A_{hsi}$ | 40 000 | 2 800 | 720 | 44 411 | 3 608 | 11 702 | 24 717 | 23 287 |
| $C_{hri}$ | 0 | 0 | 0 | 0 | 0 | 0 | 0 | 0 |
| $C_{hsi}$ | 40 000 | 2 800 | 720 | 44 411 | 3 608 | 11 702 | 24 717 | 23 287 |
| ${Sev}_{hri}$ | 0 | 0 | 0 | 0 | 0 | 0 | 0 | 0 |
| ${Sev}_{hsi}$ | 40 000 | 2 800 | 720 | 44 411 | 3 608 | 11 702 | 24 717 | 23 287 |
| $T_{hri}$ | 0 | 0 | 0 | 0 | 0 | 0 | 0 | 0 |
| $T_{hsi}$ | 0 | 0 | 0 | 0 | 0 | 0 | 0 | 0 |
| $R_{i}$ | 0 | 0 | 0 | 0 | 0 | 0 | 0 | 0 |
| $E_{1hri}$ | 0 | 0 | 0 | 0 | 0 | 0 | 0 | 0 |
| $E_{1hsi}$ | 0 | 0 | 0 | 0 | 0 | 0 | 0 | 0 |
| $S_{vi}$ | 49 182 186 | 3 434 868 | 865 614 | 53 305 512 | 4 331 829 | 14 043 978 | 29 673 408 | 27 956 829 |
| $E_{vri}$ | 0 | 0 | 0 | 0 | 0 | 0 | 0 | 0 |
| $E_{vsi}$ | 245 910 | 17 174 | 4 328 | 266 527 | 21 659 | 70 219 | 148 367 | 139 784 |
| $I_{vri}$ | 0 | 0 | 0 | 0 | 0 | 0 | 0 | 0 |
| $I_{vsi}$ | 245 910 | 17 174 | 4 328 | 266 527 | 21 659 | 70 219 | 148 367 | 139 784 |

Table 5: Description of model base parameters

| **Parameter** | **Description** | **Low transmission setting** | **High transmission setting** | **Range** | **Source** |
| --- | --- | --- | --- | --- | --- |
| $a_{i}$ | Human feeding rate per  mosquito (per day) | 0.50 | 0.50 | (0.1 – 1.0) | [21] |
| $b_{ri}$ | Probability of transmission efficiency of  resistant strain, from in-  fected vector to susceptible  human, given that contact  occurs | 0.50 | 0.50 | (0.01 – 0.27) | [21] |
| $b_{si}$ | Probability of transmission efficiency of  resistant strain, from in-  fected human to suscepti-  ble vector, given that con-  tact occurs | 0.50 | 0.50 | (0.01 – 0.27) | [21] |
| $c_{ri}$ | Probability of transmission efficiency of  resistant strain, from in-  fected human to suscepti-  ble vector, given that con-  tact occurs | Estimated from model fitting | Estimated from model fitting |  |  |
| $c_{si}$ | Probability of transmission efficiency of  sensitive strain, from in-  fected vector to human,  given that contact occurs | 0.48 | 0.48 | (0.072 - 0.64) | [21] |
| $\mu_{vi}$ | Mosquito death rate across  the entire population (per  day) | 0.033 | 0.033 | (0.0010 – 0.10) | [21] |
| $\beta_{vi}$ | Mosquito birth rate across  the entire population (per  day) | 0.13 | 0.13 | (0.020 – 0.27) | [21] |
| $\mu_{hi}$ | Human natural death rate across the en-  tire population in patch i  (per day) | Patch specific based on World Bank data | Patch specific based on World Bank data |  | [22] |
| $\beta_{hi}$ | Human birth rate across  the entire population in  patch i (per day) | Patch specific based on World Bank data | Patch specific based on World Bank data |  | [22] |
| $\gamma_{hi}$ | Rate of progression of humans from exposed state to infectious state (per day) | 1/15 | 1/15 | (0.067 – 0.20) | [12, 21] |
| $\gamma_{vi}$ | Rate of progression of mosquitoes from exposed state to infectious state (per day) | 0.083 | 0.083 | (0.029 – 0.33) | [21] |
| $\tau_{i}$ | Rate of seeking treatment  for symptomatic malaria  individuals (per day) | 1/3.5 | 1/3.5 | (1/7 – 1/3) | [23] |
| $\rho_{i}$ | Rate at which recovered  people become susceptible  again (per day) | 1/365 | 1/1825 | (1/1825 – 1/90) | [21] |
| $p_{a}$ | Proportion of asymp-  tomatic infection | 0.4 | 0.395 | (0.2 – 0.5) | [24] |
| $p_{a1}$ | Proportion of asymp-  tomatic infection after sec-  ond exposure | 0.6 | 0.7 |  | Expert opinion |
| $m_{i}$ | Mosquitoes per human (dimensionless) | 3 | 3 |  | [25] |
| $\delta_{i}$ | Human natural recovery  rate from asymptomatic  malaria individuals (per day) | 1/200 | 1/150 | (1/200 – 1/60) | [12, 23] |
| $\nu_{i}$ | Rate of progress to severe  infection (per day) | 1/7 | 1/7 | (1/10 -1/5) | [23] |
| $\zeta_{ari}$ | Relative infectiousness of  asymptomatic resistant in-  fections | 0.467 | 0.467 | (0 – 0.5) | [26, 27] |
| $\zeta_{asi}$ | Relative infectiousness of  asymptomatic sensitive in-  fections | 0.467 | 0.467 | (0 – 0.5) | [26, 27] |
| $\zeta_{tri}$ | Relative infectiousness of artemisinin partial  resistant treated infections | 0.04 | 0.04 | (0 – 0.25) | [27] |
| $\zeta_{tsi}$ | Relative infectiousness of  treated sensitive in-  fections | 0.04 | 0.04 | (0 – 0.25) | [27] |
| $\psi_{i}$ | Rate of seeking treatment  for clinical malaria individ-  uals in patch (per day) | 1/3.5 | 1/3.5 |  | [23] |
| $\eta_{si}$ | Rate of parasite clearance  for artemisinin-sensitive in-  fected individuals (per  day) | 1/3 | 1/3 | (1/3 – 1/2) | [12] |
| $\eta_{ri}$ | Rate of parasite clearance  for artemisinin-resistant in-  fected individuals (per  day) | 1/4 | 1/4 | (1/7 – 1/4) | [12] |
| $r_{ri}$ | Rate of recovery for  ACTs treated artemisinin partial  resistant infected individu-  als | 0.167 | 0.167 | (0.125 - 0.25) | [23] |
| $r_{si}$ | Rate of recovery for  ACTs treated artemisinin-  sensitive infected individu-  als | 0.167 | 0.167 | (0.125 - 0.25) | [23] |
| ${CI\_weight}_{i}$ | Local movement weight intensity |  |  |  | Estimated from model fitting |
| ${CE\_weight}_{j}$ | Foreign movement weight intensity |  |  |  | Estimated from model fitting |
| ${itn}_{use\_i}$ | Probability of sleeping un-  der a net | 0.7 | 0.7 |  | assumed |
| ${itn}_{eff\_i}$ | Effectiveness of nets in re-  ducing infectious bites | 0.40 | 0.40 | (0.897 – 0.903) | [23] |
| ${irs}_{eff\_i}$ | Effectiveness of irs in reduc-  ing infectious bites | 0.84 | 0.84 | (0.27 -0.44) | [28] |
| ${amp}_{i}$ | Amplitude of seasonal variation |  |  |  | Estimated from model fitting |
| ${peak}_{i}$ | Month of peak transmission |  |  |  | Estimated from model fitting |
| ${phase}_{i}$ | Determines when the seasonal peak occurs |  |  |  | Estimated from model fitting |
| ${PrSeekTrt}_{i}$ | Proportion of infected population reaciving treatment | 0.95 | 0.95 |  | [23] |
| $p_{i}$ | The proportion of mosquitoes surviving per day in patch $i$ |  |  |  | Estimated from model fitting |
| ${Appear}_{year}$ | Year of artemisinin partial resistance first appearance | 2025 | 2025 |  | Assumed |
| $n_{vi}$ | The parasite’s extrinsic incubation period or extrinsic cycle in days | 12 | 12 |  | [21] |

#### Human mobility

*Table 6* shows the coordinates for the eight countries in our study setting. These coordinates were used to derive a distance-weighted interaction matrix. Interactions can take place bidirectionally between two patches.

Table 6: Country coordinates

| **Subpopulation** | **Latitude** | **Longitude** | **Source** |
| --- | --- | --- | --- |
| Angola | -11.202692 | 17.873887 | [29] |
| Botswana | -22.328474 | 24.684866 | [29] |
| Eswatini | -26.522503 | 31.465866 | [29] |
| Mozambique | -18.665695 | 35.529562 | [29] |
| Namibia | -22.95764 | 18.49041 | [29] |
| South Africa | -30.559482 | 22.937506 | [29] |
| Zambia | -13.133897 | 27.849332 | [29] |
| Zimbabwe | -19.015438 | 29.154857 | [29] |

Human movement effect on malaria transmission is allowed in two ways – within and between patch movement effect. The latitude and longitude coordinates were used to estimate distance between patches and create distance matrix – symmetrical distance matrix. *Table 7* shows distance between patches with a diagonal of zeros implying zero within patch distance as assumed. The human interaction within and between patches is inversely weighted by the distance, so that the closer patches are to each other, the higher the interaction rate is relative to patches that are distant [23].

The denominator was exponentiated to $intensity weight$ as shown in *Equation (9)* [23], thus either $CI\_weight$ – local mobility intensity weight or $CE\_weight$ – foreign mobility intensity weight. Both local and foreign mobility weight intensity are assumed to be constant and inferred to parameter estimation in the transmission model. The distance between two patches is given by $d_{ij}$ which is the cartesian distance between patch $i$ and $j$ is calculated using the formula in *Equation (10)*. Movement between two patches can happen at a spatial rate $k_{ij}$ where:

$$k_{ij}=\frac{\frac{1}{1+\sqrt{d_{ij}}}}{\left( \sum_{i=1}^{8} \frac{1}{1+\sqrt{d_{ij}}} \right)^{intensity weight}}$$

(9)

$$\begin{matrix} Distance=d_{ij}=\sqrt{(}(x_{i}-x_{j})^{2}+(y_{i}-y_{j})^{2}) \end{matrix}$$

(10)

where ($x_{i}$, $y_{i}$) and ($x_{j}$, $y_{j}$) are the centroid coordinates for patches $i$ and $j$ respectively.

*Table 7* show distance matrix and movement rate matrices respectively. Then the Movement rate matrix was used in this study to mimic human movement rate within and between countries.

Table 7: Within and between patch distance matrix

|  | **Angola** | **Botswana** | **Eswatini** | **Mozambique** | **Namibia** | **South Africa** | **Zambia** | **Zimbabwe** |
| --- | --- | --- | --- | --- | --- | --- | --- | --- |
| **Angola** | 0.0000 | 1435.6073 | 2226.7584 | 2072.8144 | 1310.2043 | 2218.2257 | 1106.6007 | 1492.0740 |
| **Botswana** | 1435.6073 | 0.0000 | 830.8782 | 1202.0669 | 640.2582 | 932.6813 | 1077.1341 | 593.9420 |
| **Eswatini** | 2226.7584 | 830.8782 | 0.0000 | 969.2226 | 1370.5612 | 947.3757 | 1537.7850 | 868.7015 |
| **Mozambique** | 2072.8144 | 1202.0669 | 969.2226 | 0.0000 | 1836.2841 | 1837.6868 | 1027.2835 | 672.7382 |
| **Namibia** | 1310.2043 | 640.2582 | 1370.5612 | 1836.2841 | 0.0000 | 954.7322 | 1475.5261 | 1192.1282 |
| **South Africa** | 2218.2257 | 932.6813 | 947.3757 | 1837.6868 | 954.7322 | 0.0000 | 2005.1082 | 1430.4763 |
| **Zambia** | 1106.6007 | 1077.1341 | 1537.7850 | 1027.2835 | 1475.5261 | 2005.1082 | 0.0000 | 669.4604 |
| **Zimbabwe** | 1492.0740 | 593.9420 | 868.7015 | 672.7382 | 1192.1282 | 1430.4763 | 669.4604 | 0.0000 |

#### Seasonality

The seasonality forcing function shown in *Table 8*, was derived by normalizing the actual observed rainfall data from 2010 to 2023 for each patch. Then using the *manipulate* function, in *R*, the parameters for seasonality *Equation (11)* were determined.

*Figure 2* shows the dynamics of the seasonal forcing function used in this model. The seasonal forcing functions were modified with standardized rainfall data (2010-2023) capturing the trend or noisy characteristics of rainfall patterns of each patch.

$$\begin{matrix} \text{seasonality} & =constant+\text{amplitude}*cos\left( 2\pi\left( \frac{\text{time}}{365}-phase \right) \right)^{\text{peak}} \end{matrix}$$

(11)

Table 8: Seasonality function code for each subpopulation

| **Country** | **Code** |
| --- | --- |
| Angola | $0.41 + 0.40 * cos\left( 2\pi\left( \frac{\text{time}}{365}-0.07 \right) \right)^{\text{1}}$ |
| Botswana | $0.24 + 0.24 * cos\left( 2\pi\left( \frac{\text{time}}{365}-0.07 \right) \right)^{\text{1}}$ |
| Eswatini | $0.18 + 0.18 * cos\left( 2\pi\left( \frac{\text{time}}{365}-0.05 \right) \right)^{\text{1}}$ |
| Mozambique | $0.43+ 0.41* cos\left( 2\pi\left( \frac{\text{time}}{365}-0.13 \right) \right)^{\text{1}}$ |
| Namibia | $0.27 + 0.26 * cos\left( 2\pi\left( \frac{\text{time}}{365}-0.08 \right) \right)^{\text{1}}$ |
| South Africa | $0.33 + 0.27 * cos\left( 2\pi\left( \frac{\text{time}}{365}-0.07 \right) \right)^{\text{1}}$ |
| Zambia | $0.41 + 0.40 * cos\left( 2\pi\left( \frac{\text{time}}{365}-0.07 \right) \right)^{\text{1}}$ |
| Zimbabwe | $0.29 + 0.28 * cos\left( 2\pi\left( \frac{\text{time}}{365}-0.10 \right) \right)^{\text{1}}$ |

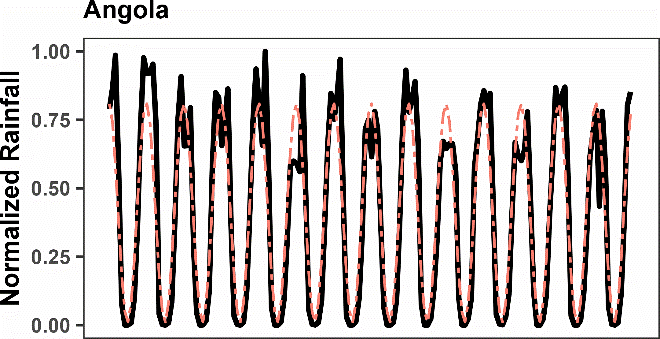

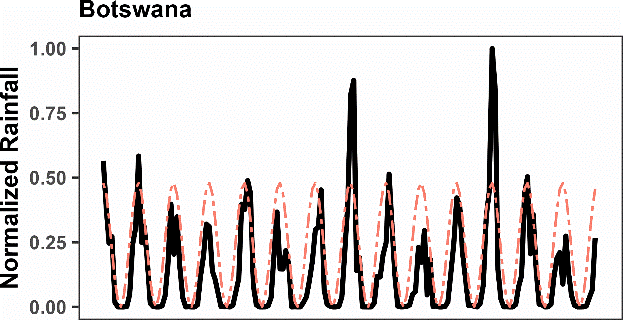

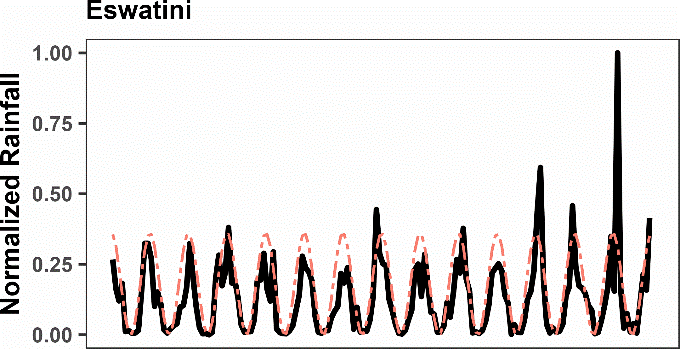

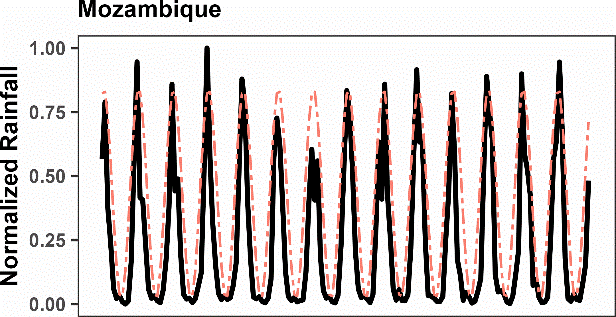

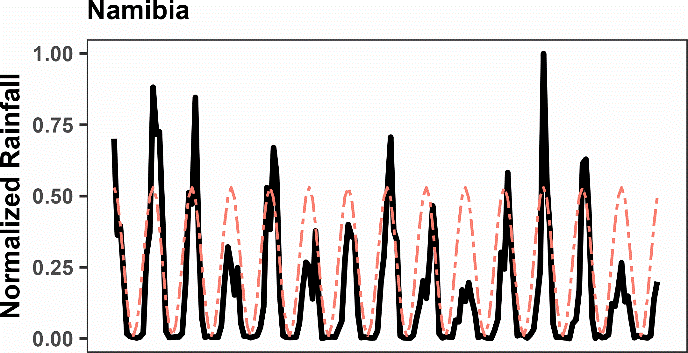

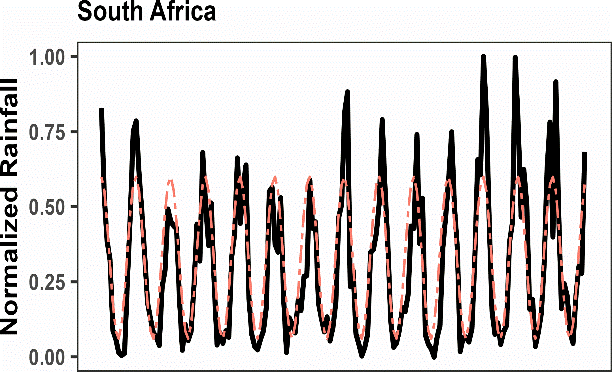

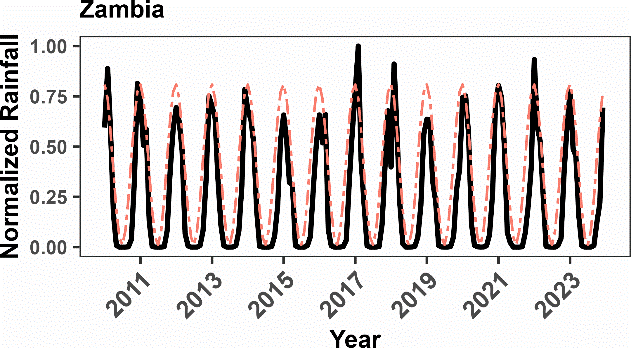

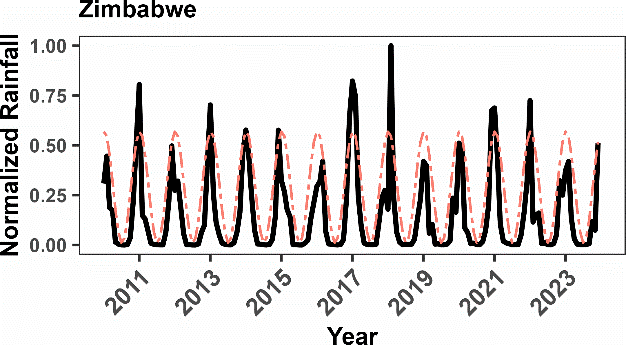

Figure 2: Seasonality forcing function based on monthly rainfall data in each country. Black solid line is the observed normalized rainfall (2010-2023) while salmon two dashed line is the fitted normalised rainfall.

#### Model assumptions

To maintain simplicity and interpretability, we made several assumptions based on existing biological epidemiological data:

1. In the absence of vector control, the human population is equally attractive to mosquito biting
2. It is assumed that odyssean malaria does not occur
3. First line antimalarials in use are ACTs in all subpopulations
4. Full susceptibility to the partner drug is assumed
5. Interaction between patches does not lead to a change in permanent place of residence
6. Individuals in each patch are well mixed and homogeneous except for disease status and strain
7. Only the human population can recover from malaria
8. Mosquito longevity in the absence of vector control is independent of malaria infection
9. No coinfection, thus no infection by both strains at the same time
10. Distance between people within the same country is assumed to be zero

#### Model data fitting and model validation

We split our data extracted from the WHO annexes from 2015 to 2024 into two independent data sets: the model fitting (2015 - 2020) and the model validation (2021 - 2024) datasets. The extracted data were the reported *P. falciparum* annual cases. Model fitting was performed using the model fitting dataset [23] adjusted for routine IRS and ITN coverage between 2015 and 2020 and connectivity between patches. Vector control coverage data, routine IRS and ITN for each country, from 2000 to 2024 was extracted from Malaria Atlas Project (MAP) [30]. The model is run from year 2000 for 1 825 days before introducing vector control and then allowed to run for 10 years more before being fitted to *P. falciparum* annual reported cases data from 2015 to 2020 . Data fitting was performed using maximum likelihood estimation (MLE) assuming the Poisson distribution with parameter, $\lambda$, equal to the annual *P. falciparum* reported cases. Sheetal eta al. (2015) defined the parameter $\lambda$ as the number of treated cases per week [23].

The optimised parameters were the probability of mosquito survival, local movement weight intensity, foreign movement weight intensity and mosquito-to-human ratio parameters. The process was carried out to reduce the uncertainty of the parameters to obtain high credibility of the model [31]. The *R* software [32] *OptimParallel* function was used for the model calibration with 1000 iterations.

The model was then run with the fitted parameters for another five years (2021 to 2024) and compared with WHO *P. falciparum* annual reported cases data from 2021 and 2024 with respective 95% confidence interval for uncertainty. This is the most reliable and preferred way to validate model simulations [33]. However, given that there was no data on artemisinin partial resistant cases as of 2024 in our study setting[10], sensitivity analysis was carried out to support model validation [33, 34].

#### Model scenarios

*Table 9* shows study scenarios in which artemisinin partial resistant infections may enter circulation and spread either low transmission, high transmission or multinational. All these scenarios were then compared to the baseline scenario - artemisinin partial resistance free scenario.

Table 9: Description of study scenarios

| **Scenario** | **Gametocyte Carriage Ratio 3 days after ACTs treatment (Artemisinin Partial Resistant: Wild type)** | | **Time to Parasite Clearance (Days)** | **Artemisinin Partial Resistant Mutation** | | **Source** |
| --- | --- | --- | --- | --- | --- | --- |
| Status quo (Artemisinin-resistant free) | - | ≤ 3 days | | |  | [35] |
| First appear in Botswana | 2:1 | ≥ 4 days | | | *Pfkelch13* | [13, 35, 36] |
| First appear in Angola | 2:1 | ≥ 4 days | | | *Pfkelch13* | [13, 35, 36] |
| Multi-country introduction | 2:1 | ≥ 4 days | | | *Pfkelch13* | [13, 35, 36] |

The artemisinin partial resistant free scenario is the baseline (status quo) where all the treated in each country received a 3-day course of ACT and assumed to fully comply with the full course of treatment. Traditional treatments and/or drugs were not incorporated. The second, third and fourth scenarios are described in terms of first entrance into circulation of artemisinin partial resistance. First appearance could be in low transmission patch, high transmission patch or multi-patch. The scenarios were born out of the randomness in first entrance into circulation and ultimate spread in connected countries. An initial seed of two artemisinin partial resistant infectious humans and mosquitoes were hypothetically introduced in each patch at day 9126 after running the model for 25 years (2000 to 2024) and then allowed to run for 15 more years (2026 to 2039).

#### Sensitivity analysis

We determined the importance of various risk factors responsible for the spread of artemisinin partial resistant malaria infections by evaluating the impact of the variations of each parameter of the model [37] . We calculated partial rank correlation coefficients (PRCCs) using Latin Hypercube Sampling (LHS)[37] that is described by Hoare et al. (2008) as an efficient method for sampling multi-dimensional parameter space [38].

The PRCC value, that lies within -1 to +1 interval, was used to describe the contribution of each parameter to the artemisinin partial resistant cases. Positive (negative) PRCC imply a positive (negative) correlation between the respective model parameter and the outcomes [37]. A positive (negative) change in parameters in question will increase(reduce) the outcome values [39]. The bigger the absolute PRCC value the more sensitive the model outcome is the parameter [37]. The model outputs we considered were the simulated artemisinin partial resistant cases and the total (sum of artemisinin partial resistant and sensitive cases) number of cases.

A probability distribution was assigned to each parameter as suggested by Hoare et al (2008) [38, 40]. Uncertainty regarding all parameters bound between 0 and 1 was represented by the beta distribution while gamma distribution was used to represent uncertainty regarding all parameters that can take any positive value and a total of 100 computations of each output variable of interest for each country were generated [37]. The tornado plot for PRCC was used to indicate each parameter’s uncertainty in contributing to the variability in time for the spread of artemisinin partial resistance [38]. *R* software [32] was used for all simulations of the models, data analyses, visualizations and all tests were done at 5% level of significance.

### Results

*Table 10* shows the parameters obtained through data fitting and *Figure 3* shows model validation for the eight SADC-E8 countries.

Table 10: Optimised model parameters

| **Country** | $\boldsymbol{m}_{\boldsymbol{i}}$ | $\mu_{vi}$ | $\boldsymbol{p}_{\boldsymbol{i}}$ | ${CI\_weight}_{i}$ | ${CE\_weight}_{j}$ |
| --- | --- | --- | --- | --- | --- |
| Angola | 3 | 0.140 | 0.869 | 1.000 | 0.20 |
| Botswana | 3 | 0.646 | 0.524 | 1.000 | 0.30 |
| Eswatini | 3 | 0.522 | 0.593 | 1.000 | 0.32 |
| Mozambique | 3 | 0.095 | 0.909 | 1.000 | 0.10 |
| Namibia | 3 | 0.454 | 0.635 | 1.000 | 0.38 |
| South Africa | 3 | 0.579 | 0.560 | 1.000 | 0.38 |
| Zambia | 3 | 0.060 | 0.942 | 1.000 | 0.10 |
| Zimbabwe | 3 | 0.330 | 0.719 | 1.000 | 0.20 |

***Abbreviations:*** $m_{i}$ *is the relative density of mosquitoes in relation to human density,*
$\mu_{vi}$ *is the mosquito death rate per day,*
$p_{i}$ *is the proportion of mosquitoes surviving per day,*
${CI\_weight}_{i}$ *is the local mobility intensity weight and*
${CE\_weight}_{j}$ *is the external mobility intensity weight.*

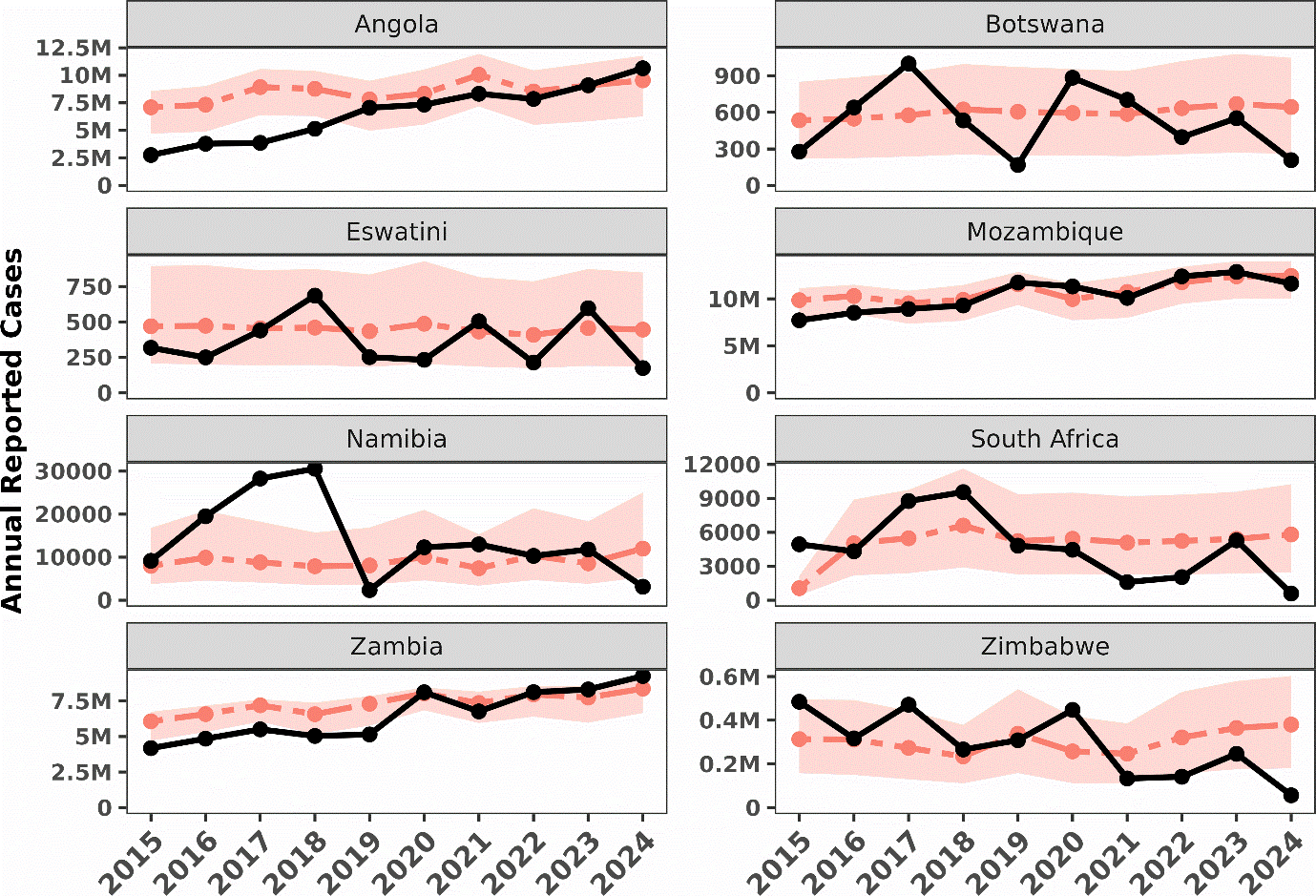

Figure 3: Model data fitting(2015-2020) and validation (2021 – 2024) using annual P. falciparum malaria reported cases. The solid black line is the observed while the two dashed salmon line is the simulated annual P. falciparum malaria reported cases.The lightly salmon coloured region represents 95% uncertainty range for annual case predictions.

*Figure 4* shows the exponential rise in daily new artemisinin partial resistant infections, over time, as a proportion of total daily new infections in our worst scenario – multipatch hypothyetical introduction of artemisinin partial resistance. However, with relatively small increase annual reported cases overall.

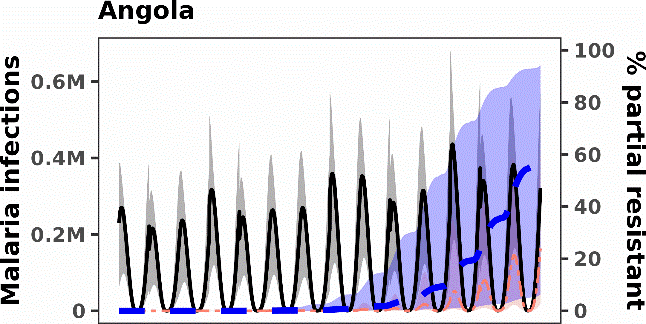

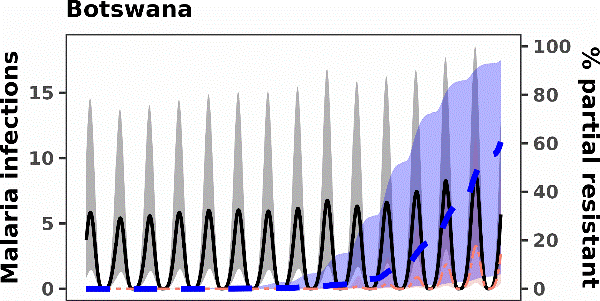

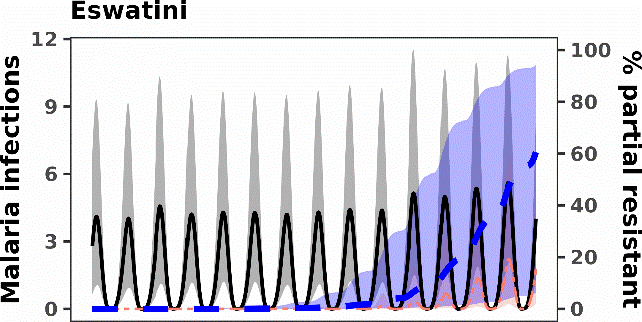

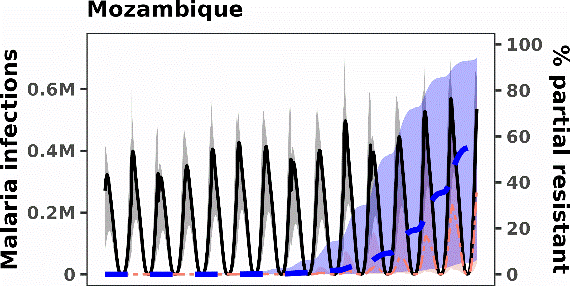

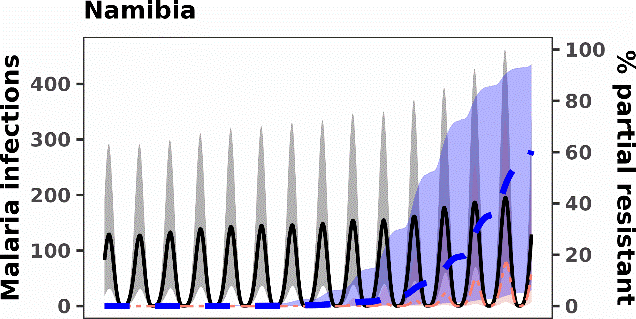

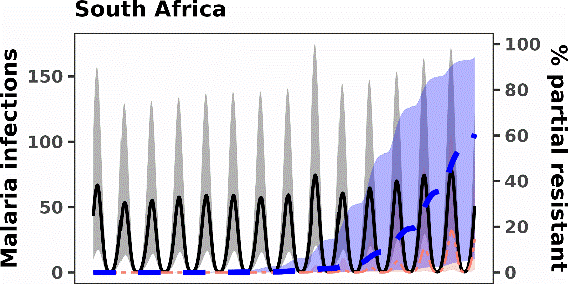

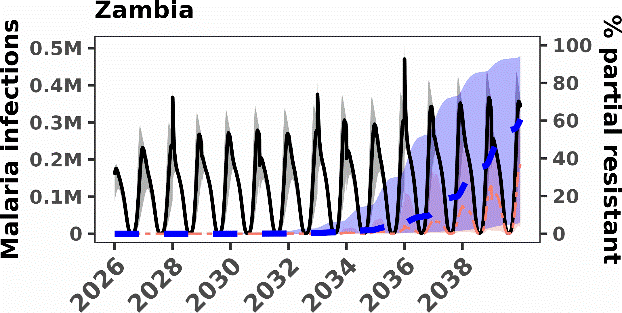

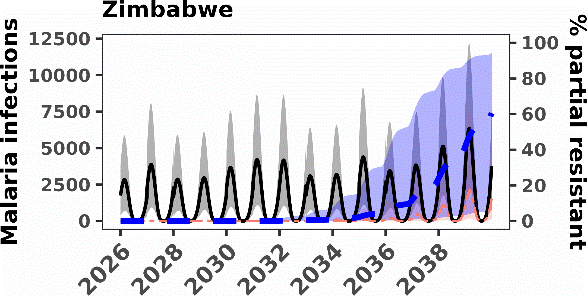

Figure 4: Malaria daily new infections from 2026 to 2039 after artemisinin partial resistance was artificially introduced in all connected (assuming status quo calibrated connectivity level) patches at the same time. The black solid line is the total malaria new infections; the salmon line is the artemisinin partial resistant new infections while the blue dashed line is the artemisinin partial resistant new infections as a percentage of total malaria new infections, lightly shaded is the 95% uncertainty region.

#### Sensitivity analysis

Sensitivity analysis results shown in *Table 11* are for the parameters signifcantly (p-value <0.05) correlated to the number of malaria cases attributable to artemisinin partial resistant infections and the respective PRCC value and the 95% CI. *Figure 5* shows the most sensitive parameters and respective direction of influence on the spread of artemisinin partial resistance.

Table 11: Artemisinin partial resistant cases PRCC results with p-values and 95% confidence intervals (CI) for significant parameters.

| **Country** | **Parameter** | **PRCC** | **95% CI** | **P-value** |
| --- | --- | --- | --- | --- |
| Angola | Probability of transmission from mosquito to human | 0.970 | 0.956 – 0.980 | <0.01 |
|  | Probability of transmission from human to mosquito | 0.831 | 0.759 – 0.884 | <0.01 |
|  | Daily biting rate per mosquito | 0.716 | 0.605 – 0.800 | <0.01 |
| Botswana | External human mobility intensity weight | 0.992 | 0.988 – 0.994 | <0.01 |
|  | Daily biting rate per mosquito | 0.856 | 0.793 – 0.901 | <0.01 |
|  | Probability of transmission from mosquito to human | 0.247 | 0.053 – 0.423 | 0.013 |
| Eswatini | External human mobility intensity weight | 0.986 | 0.979 – 0.991 | <0.01 |
|  | Daily biting rate per mosquito | 0.831 | 0.758 – 0.883 | <0.01 |
| Mozambique | Probability of transmission from mosquito to human | 0.964 | 0.948 – 0.976 | <0.01 |
|  | Probability of transmission from human to mosquito | 0.783 | 0.694 – 0.849 | <0.01 |
|  | Daily biting rate per mosquito | 0.627 | 0.491 – 0.733 | <0.01 |
| Namibia | External human mobility intensity weight | 0.987 | 0.980 -0.991 | <0.01 |
|  | Daily biting rate per mosquito | 0.871 | 0.813 – 0.911 | <0.01 |
|  | Probability of transmission from mosquito to human | 0.294 | 0.105 – 0.464 | 0.003 |
| South Africa | External human mobility intensity weight | 0.990 | 0.985 -0.993 | <0.01 |
|  | Daily biting rate per mosquito | 0.878 | 0.824 – 0.917 | <0.01 |
|  | Probability of transmission from mosquito to human | 0.207 | 0.011 – 0.388 | 0.039 |
| Zambia | Probability of transmission from mosquito to human | 0.925 | 0.891 – 0.949 | <0.01 |
|  | Probability of transmission from human to mosquito | 0.640 | 0.508 – 0.743 | <0.01 |
|  | Daily biting rate per mosquito | 0.631 | 0.496 – 0.736 | <0.01 |
| Zimbabwe | Probability of transmission from mosquito to human | 0.912 | 0.872 – 0.940 | <0.01 |
|  | Daily biting rate per mosquito | 0.642 | 0.509 – 0.744 | <0.01 |
|  | Probability of transmission from human to mosquito | 0.633 | 0.499 – 0.738 | <0.01 |

*CI = Confidence interval, * PRCC= Partial rank correlation coefficients

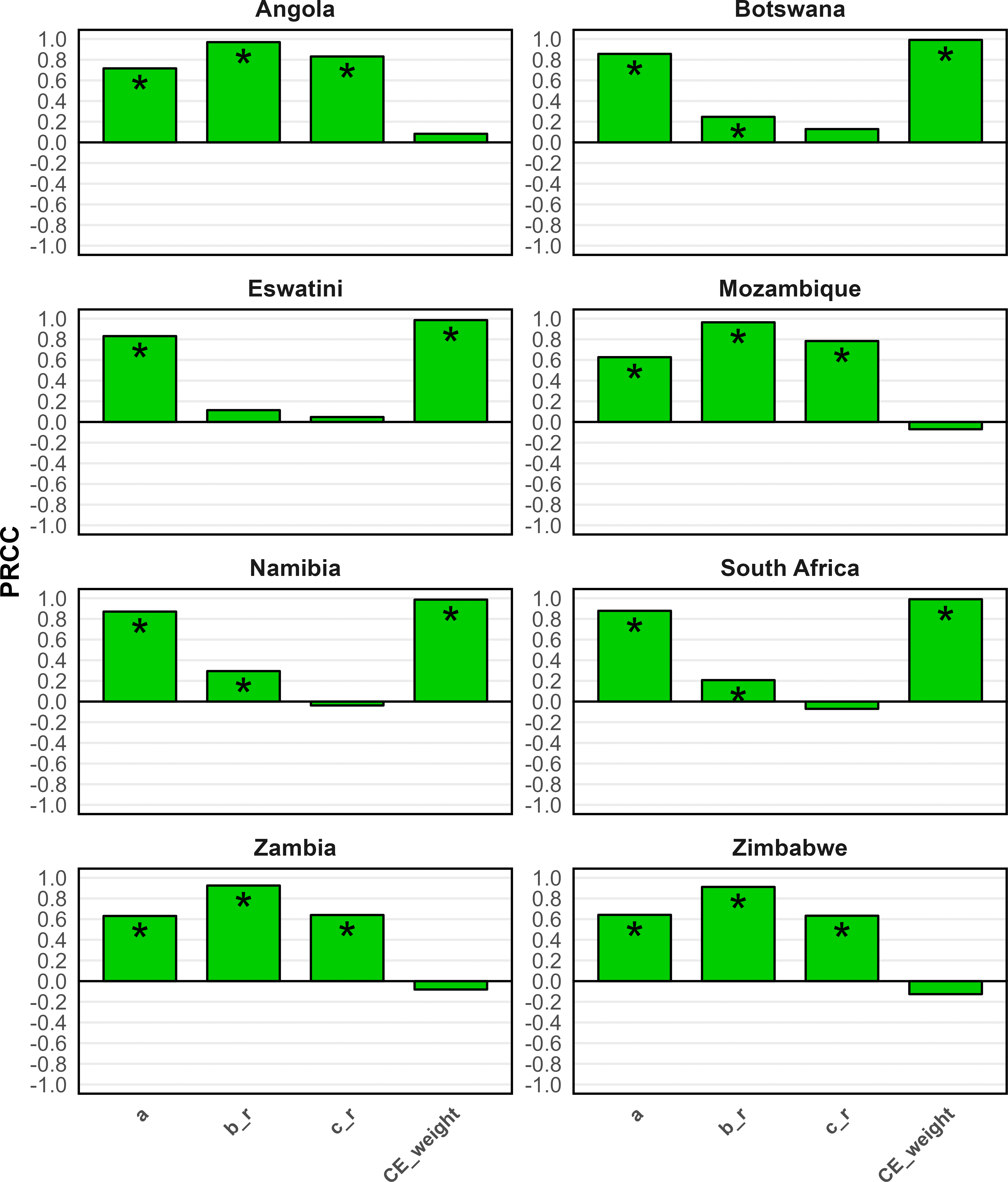

Figure 5: PRCC values showing the sensitivities of model output (reported resistant cases) with respect to the input parameters $a_{i}$(biting rate per mosquito per day), $b_{ri}$(probability of transmission from infectious mosquito to human), $c_{ri}$(probability of transmission from infectious human to mosquito), and ${CE\_weight}_{j}$(human movement intensity weight between countries). A * symbol is used to indicate significant parameters.
