## Supplementary figures and images for "Borderless battles: Modelling the spread of artemisinin partial resistance in connected subpopulations in southern Africa"

### S2 Fig 1

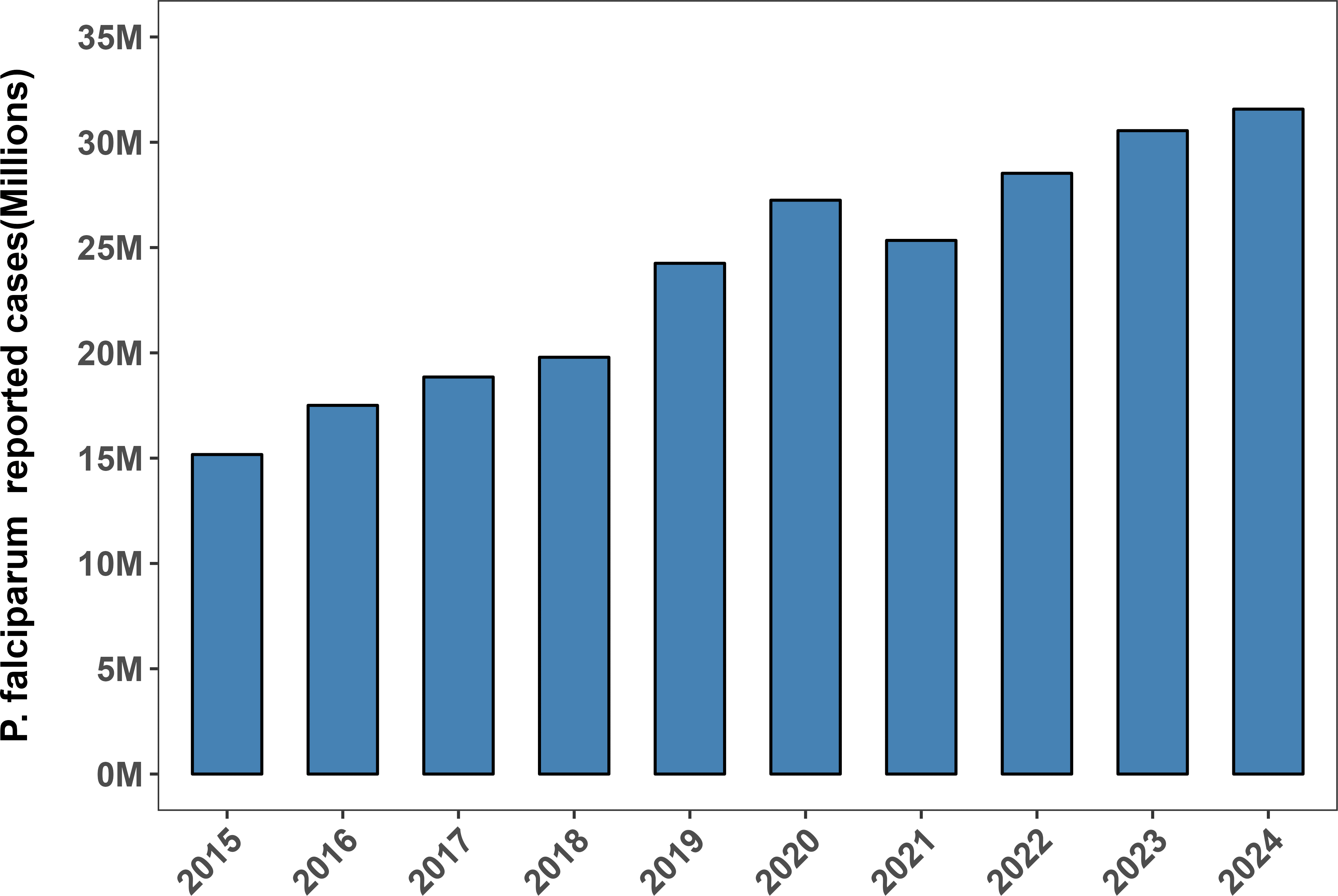

### S2 Fig 2a

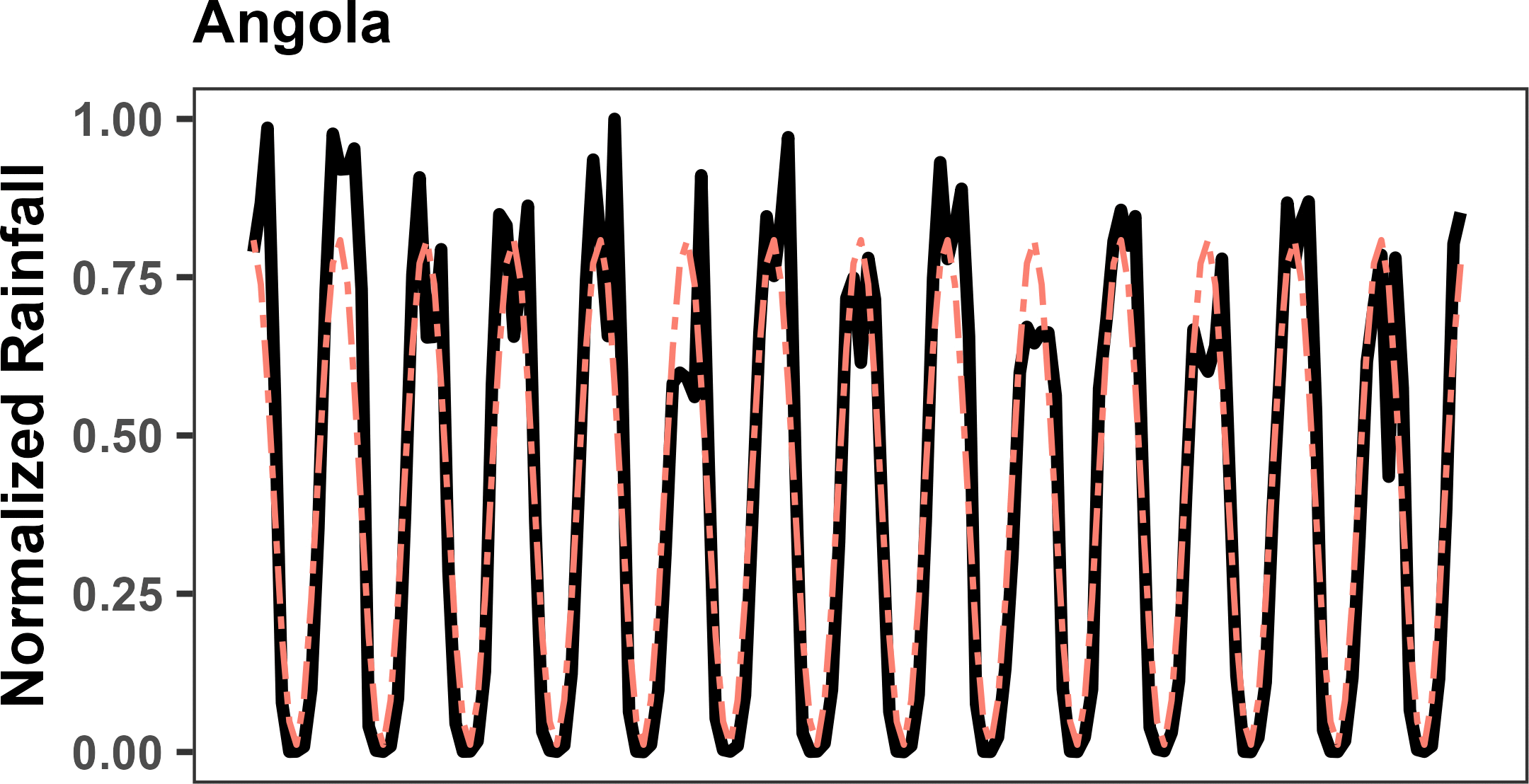

### S2 Fig 2b

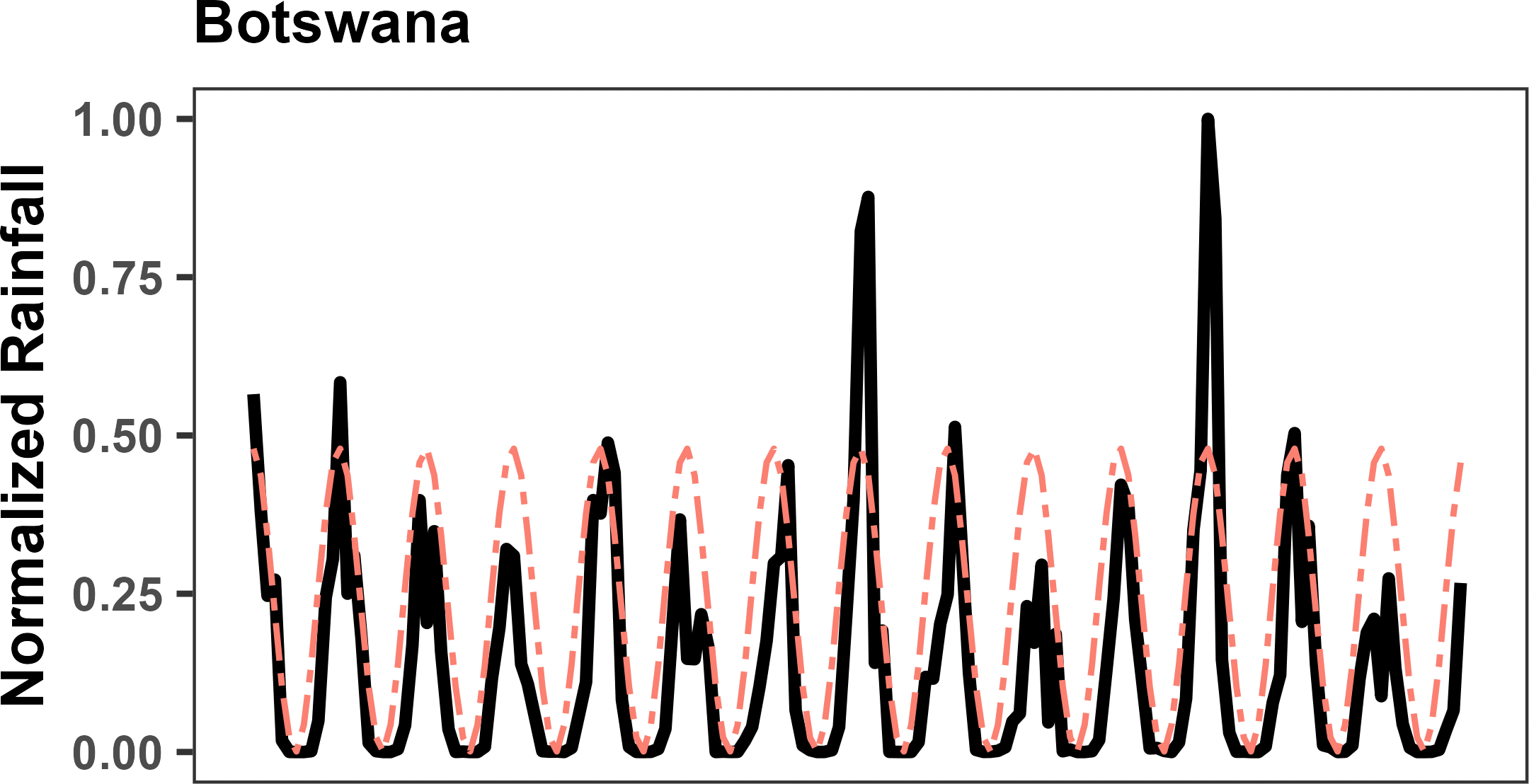

### S2 Fig 2c

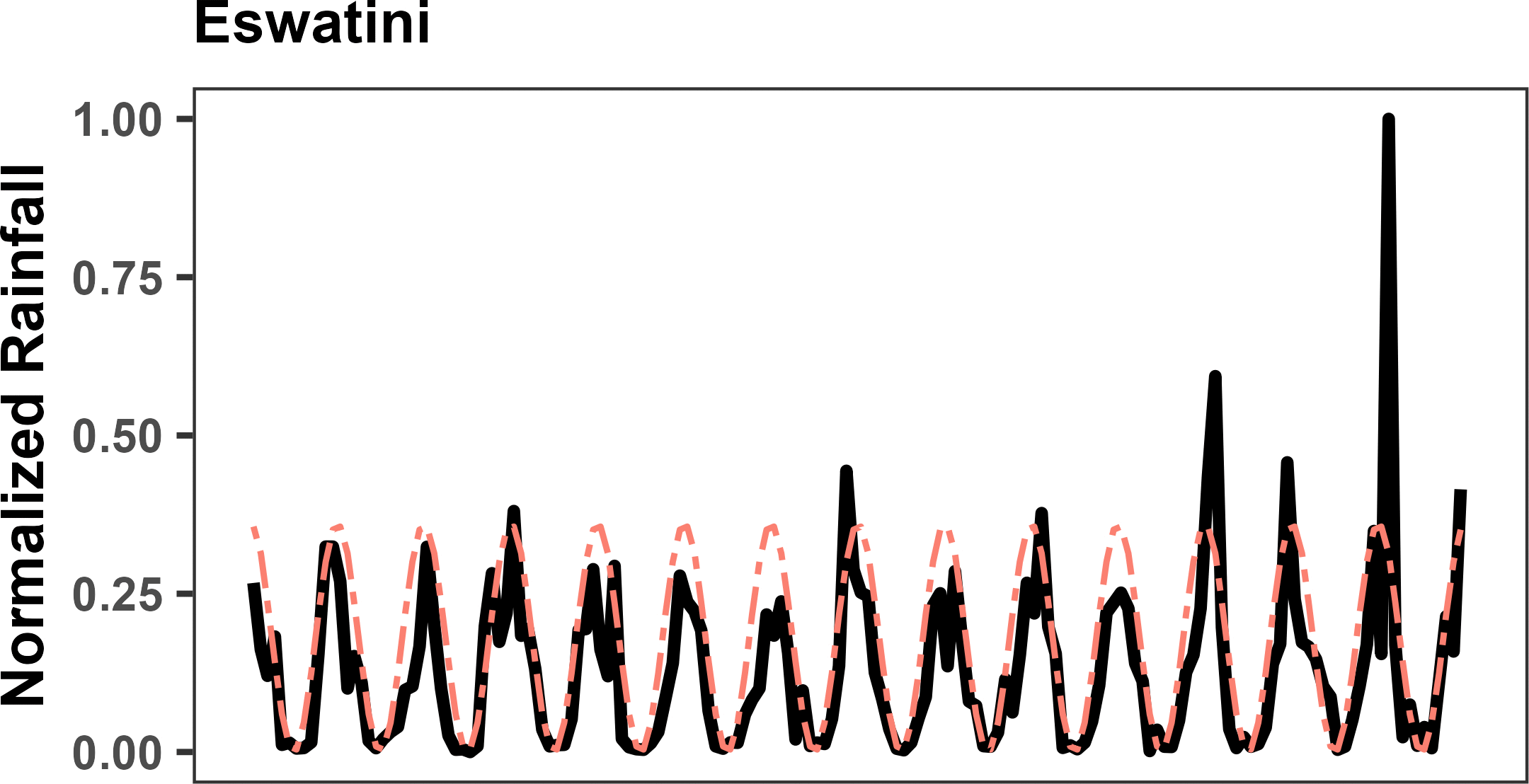

### S2 Fig 2d

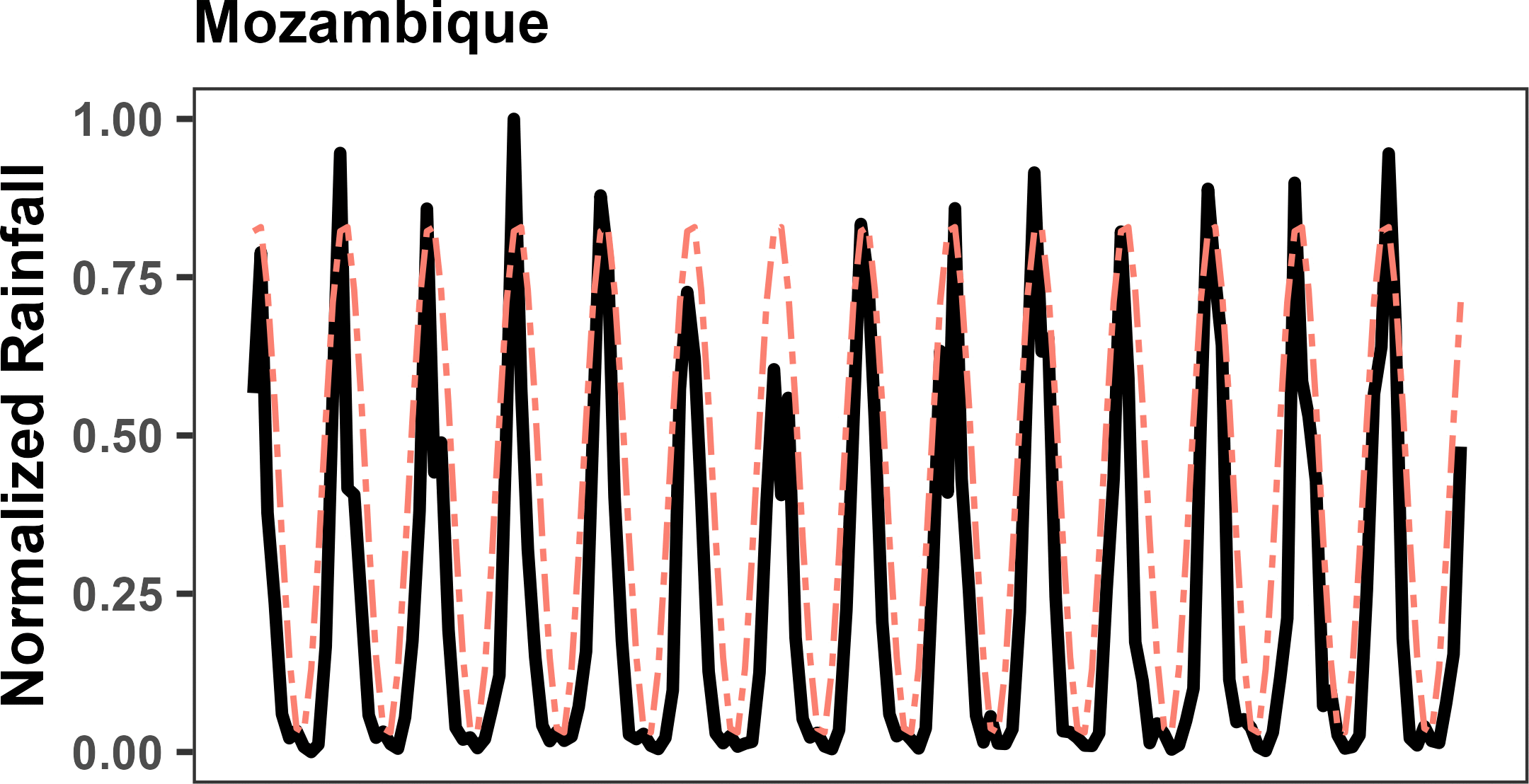

### S2 Fig 2e

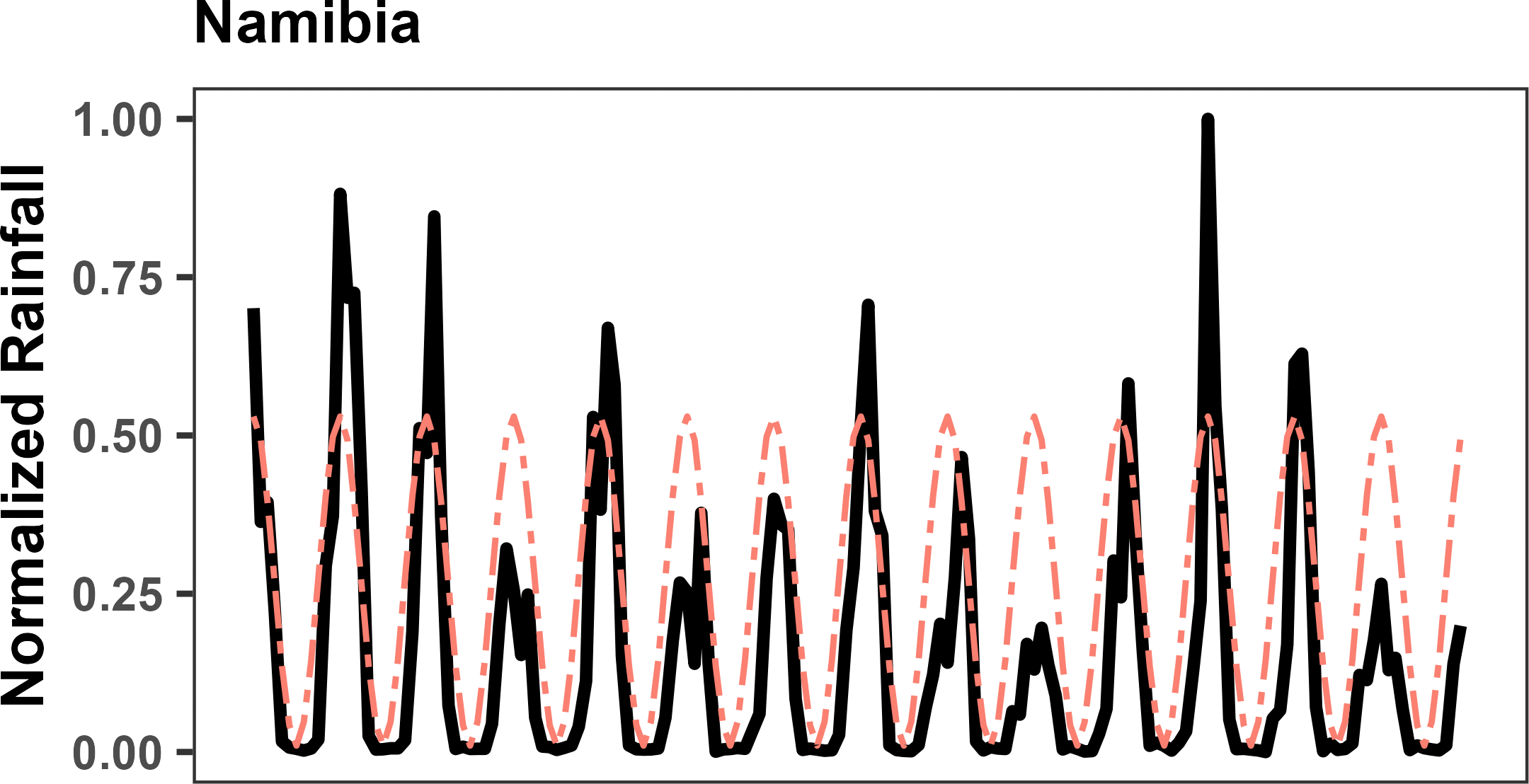

### S2 Fig 2f

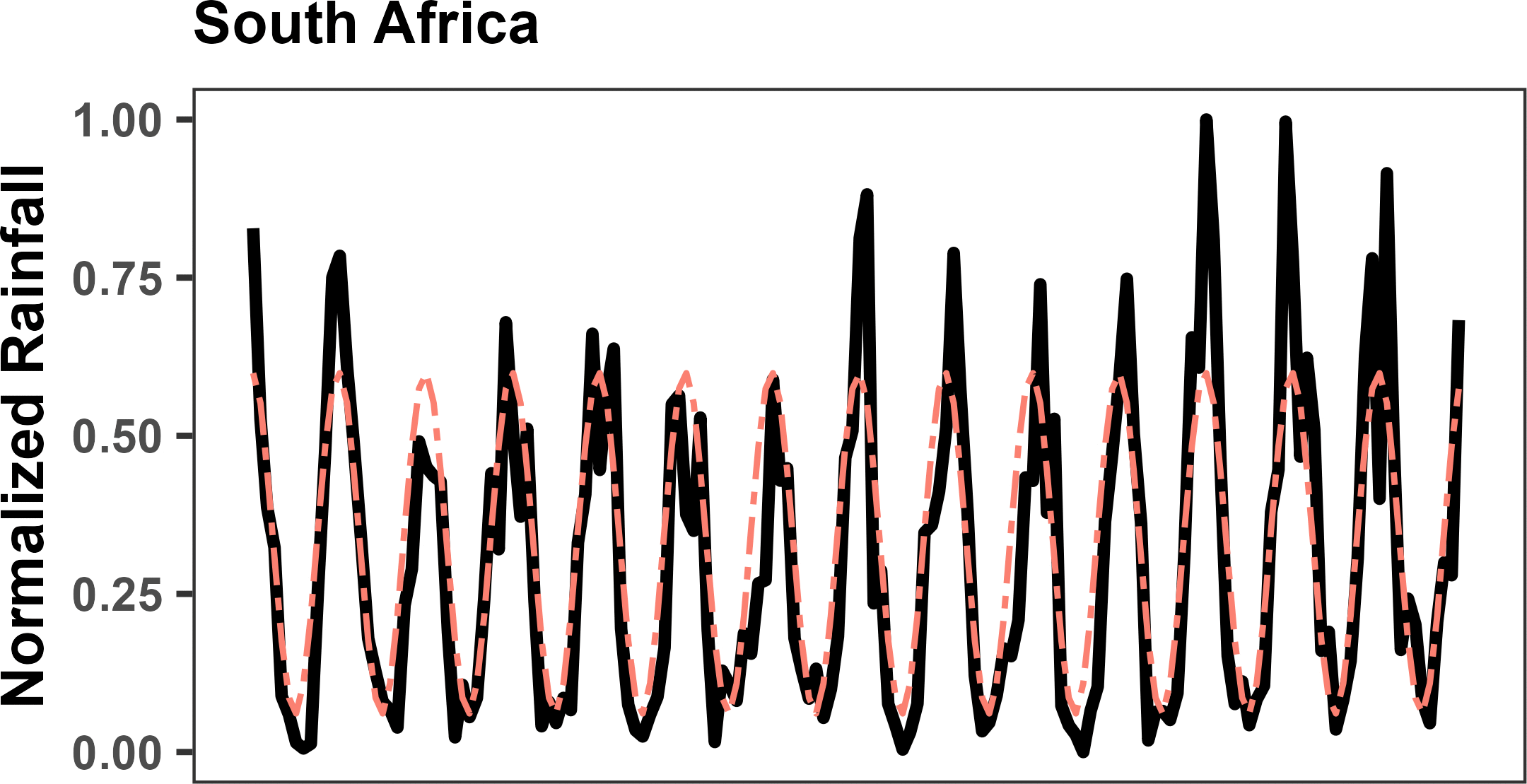

### S2 Fig 2g

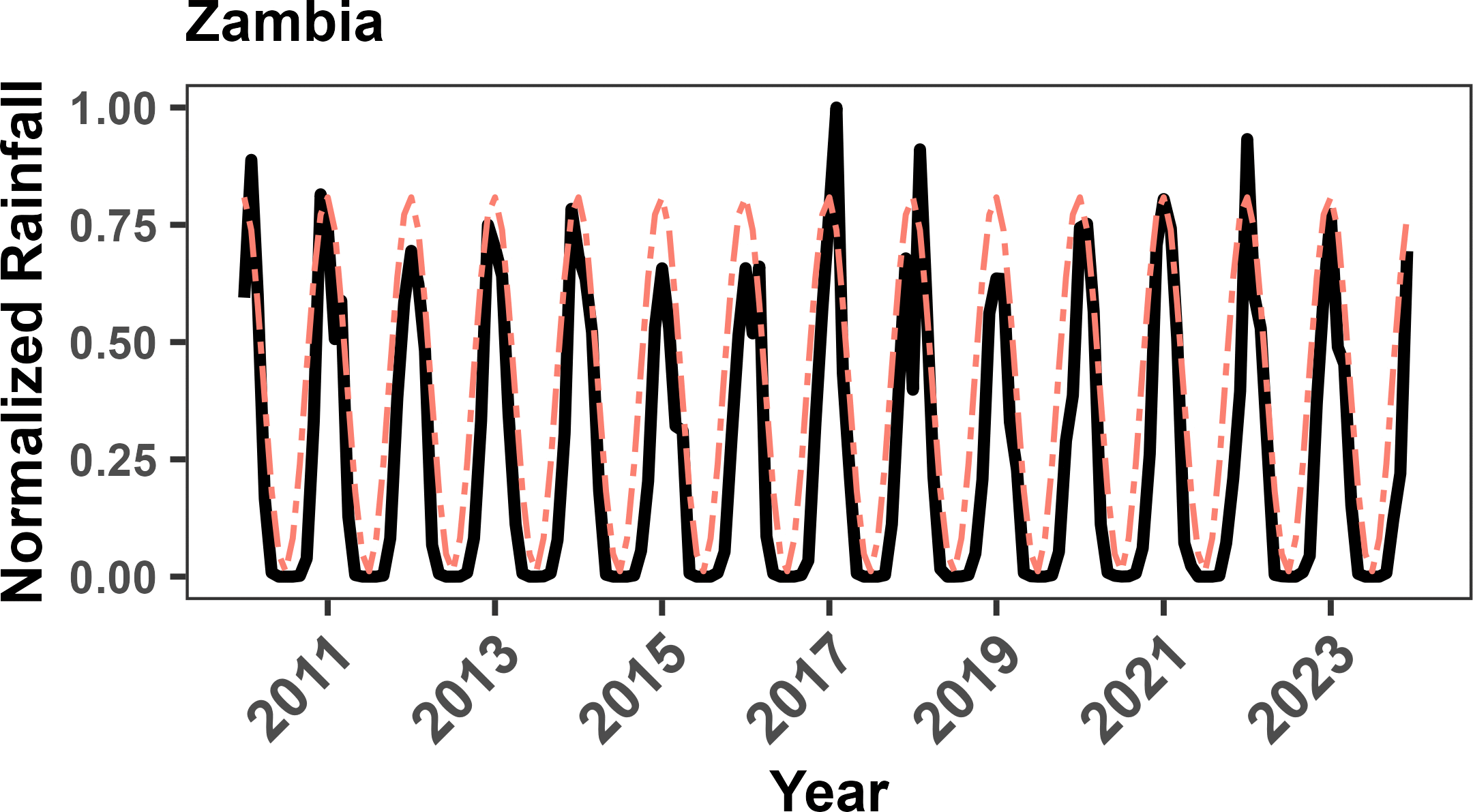

### S2 Fig 2h

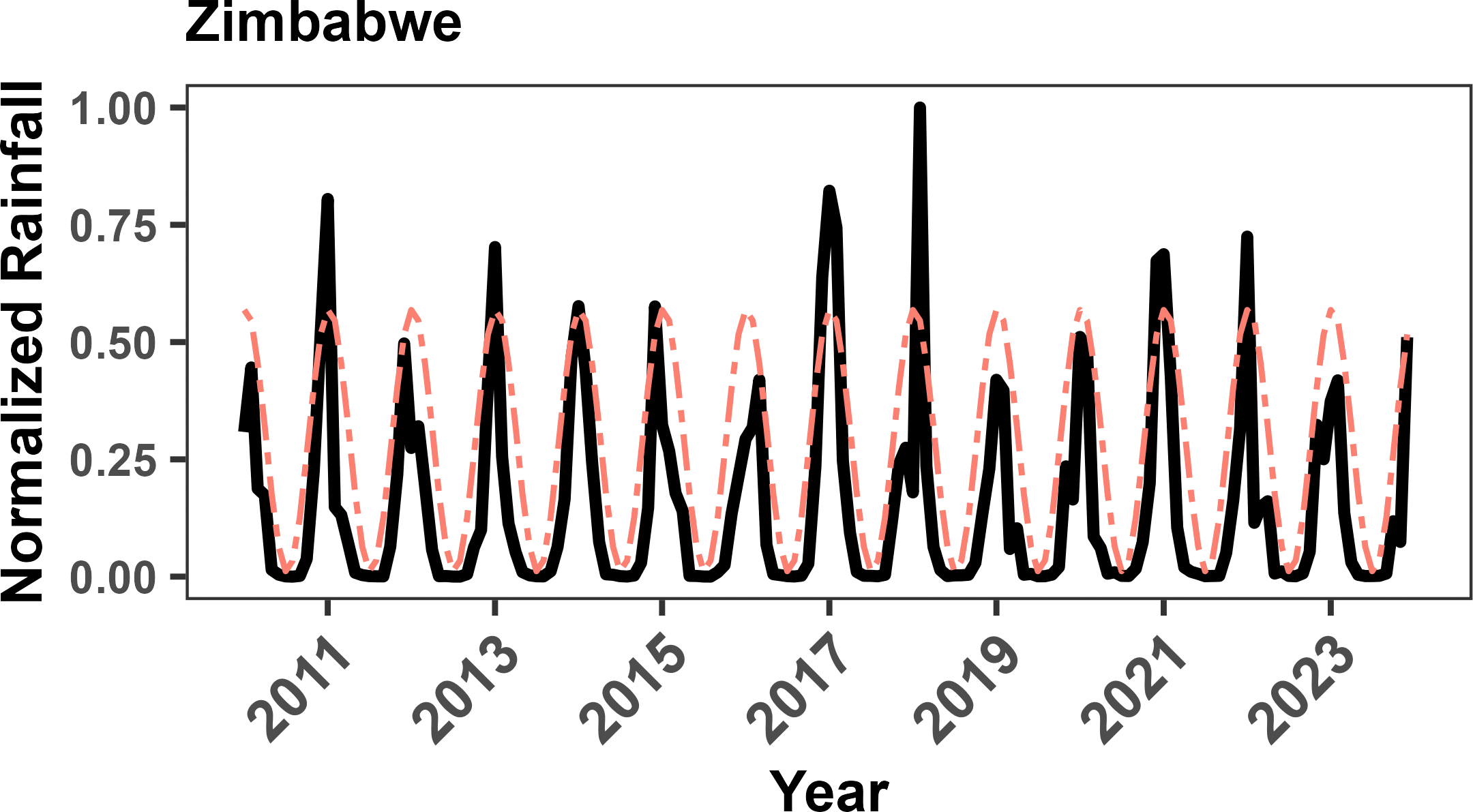

### S3 Fig 3

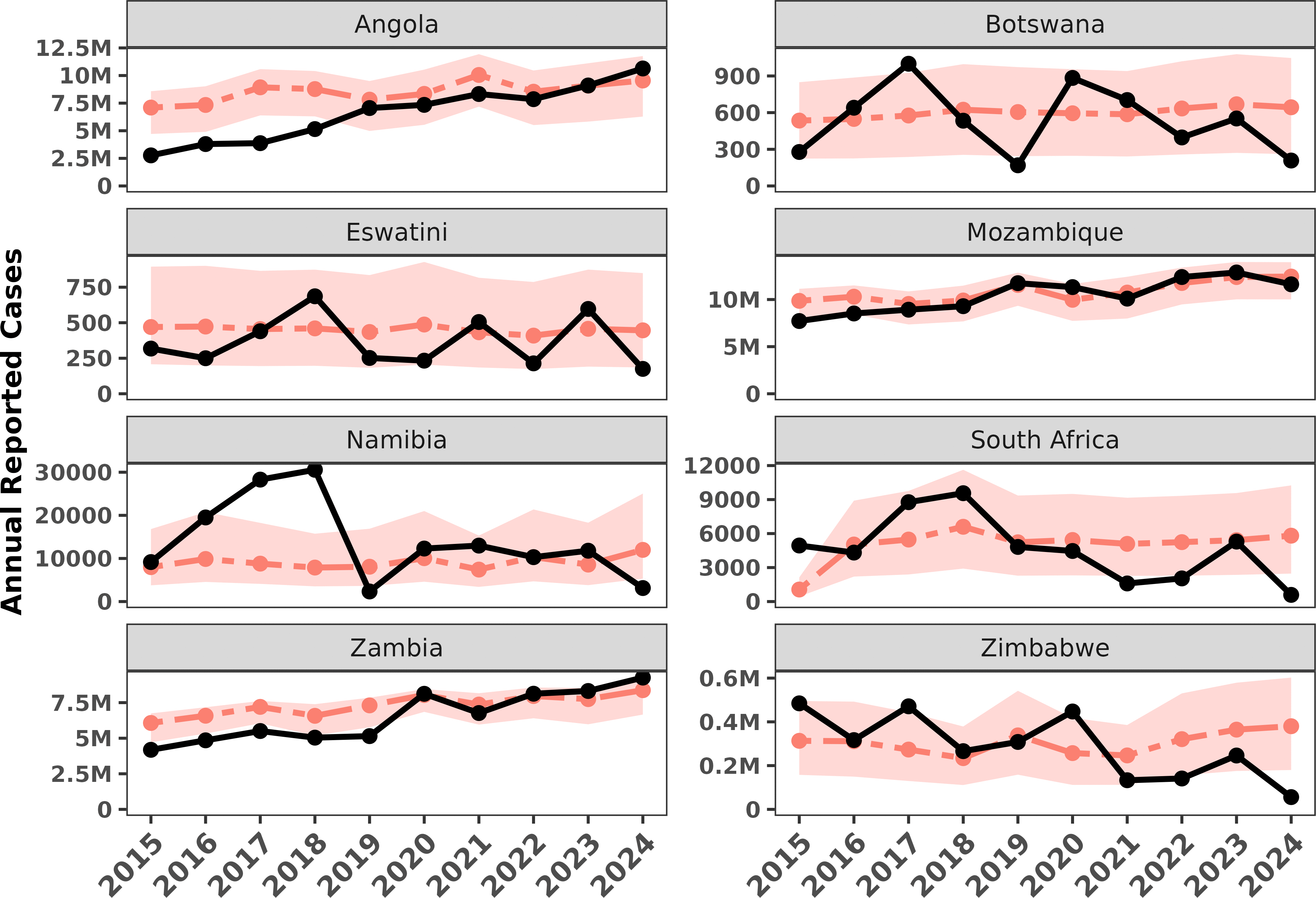

### S4 Fig 4a

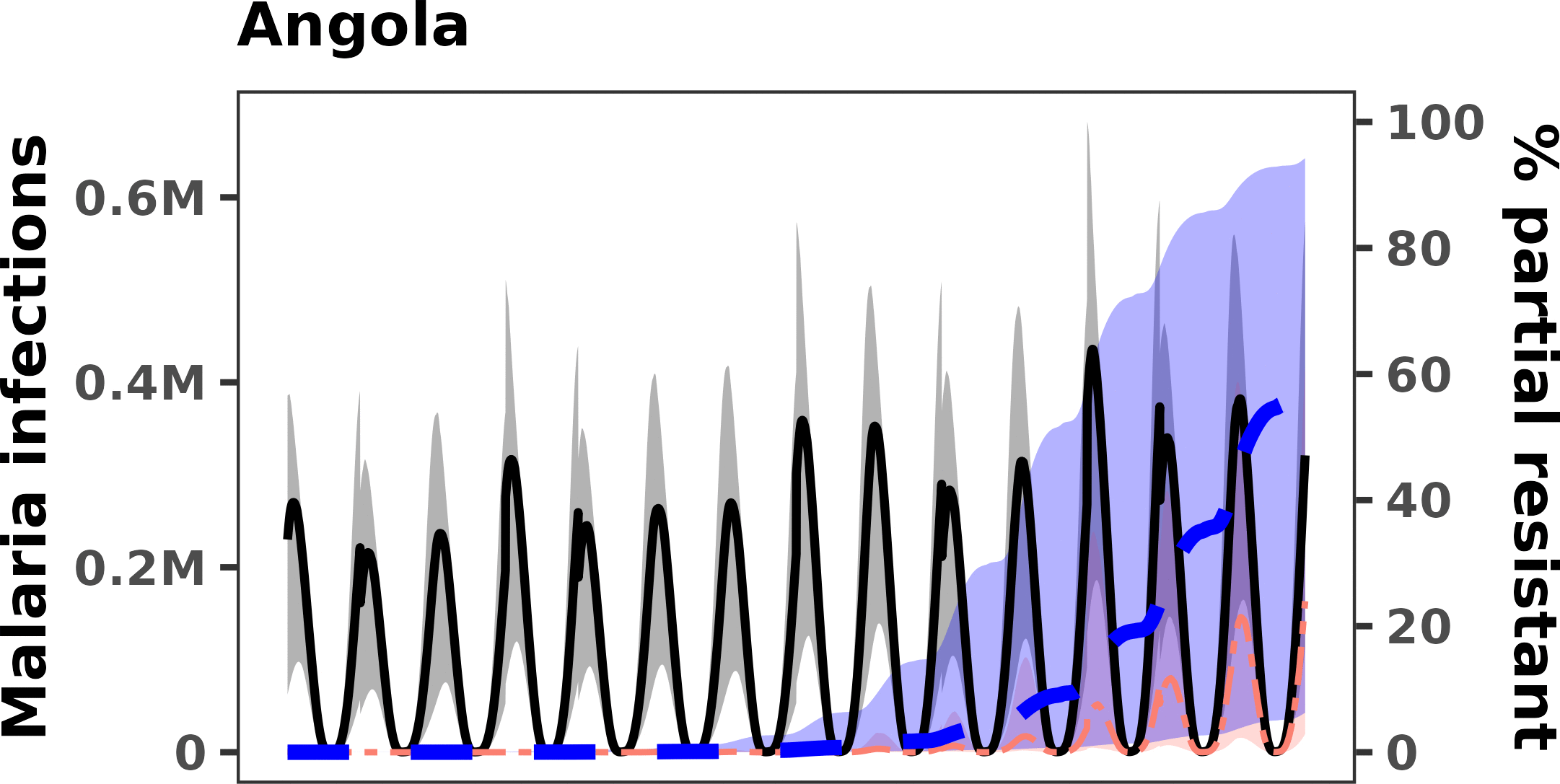

### S4 Fig 4b

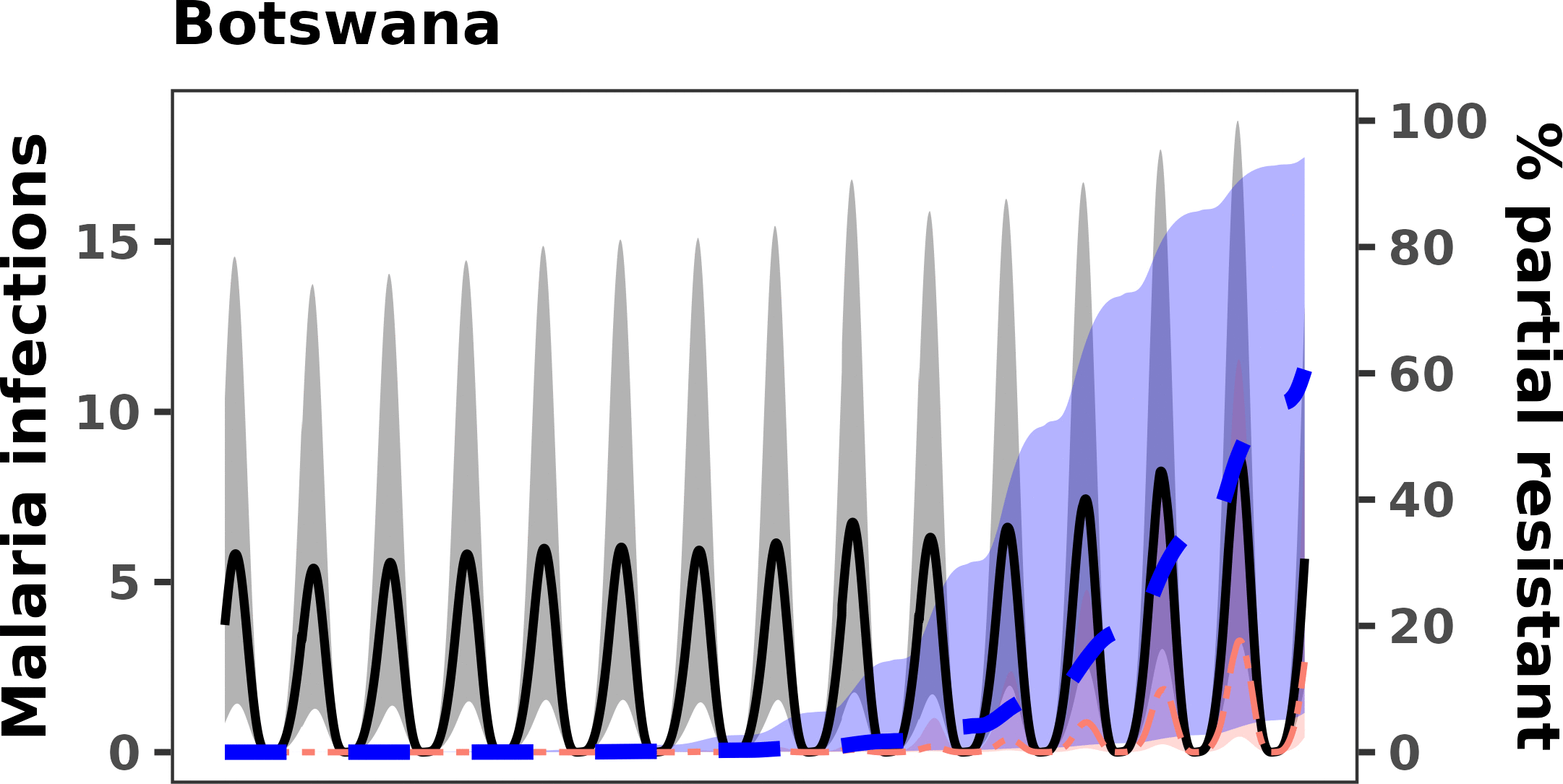

### S4 Fig 4c

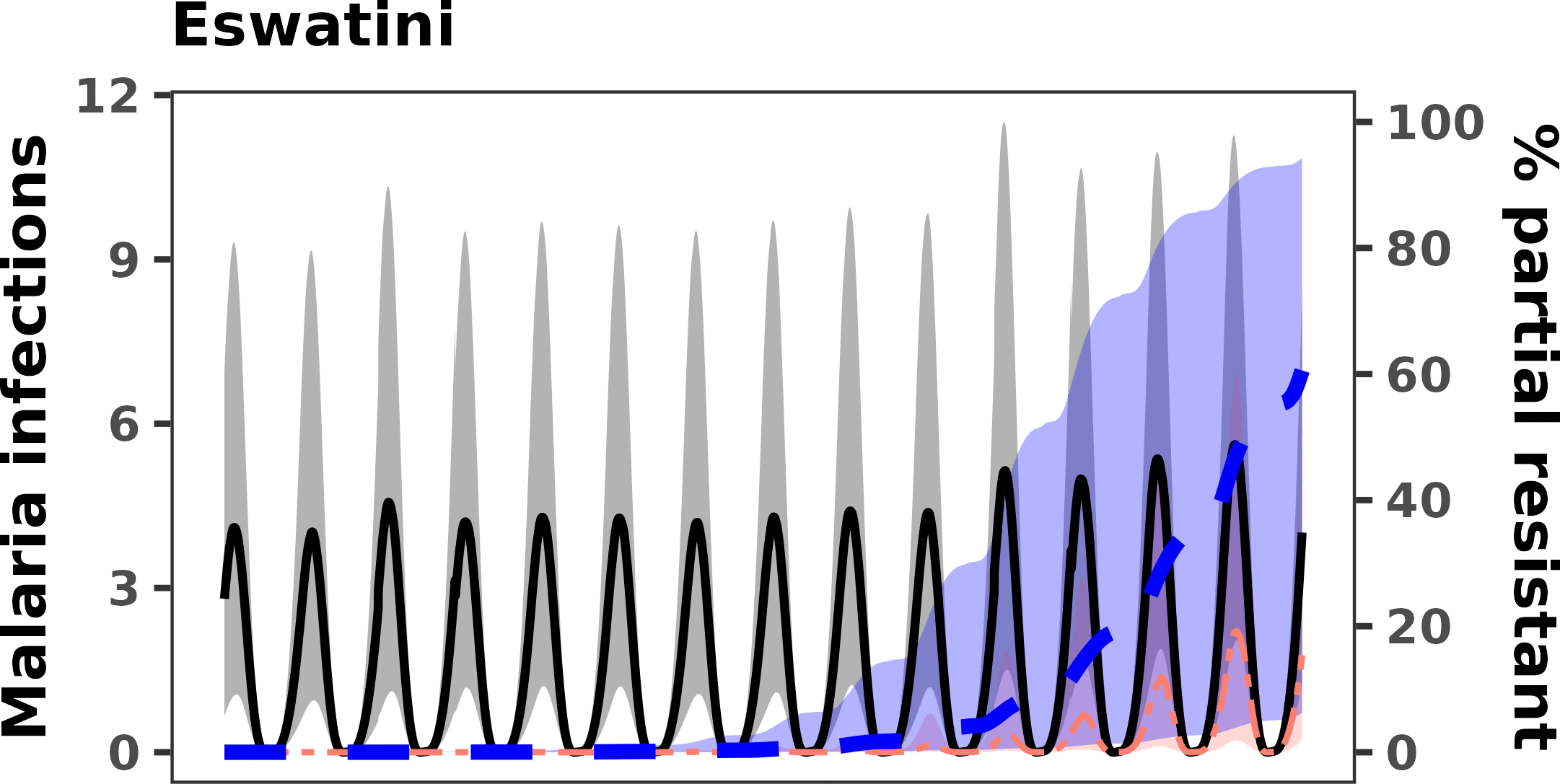

### S4 Fig 4d

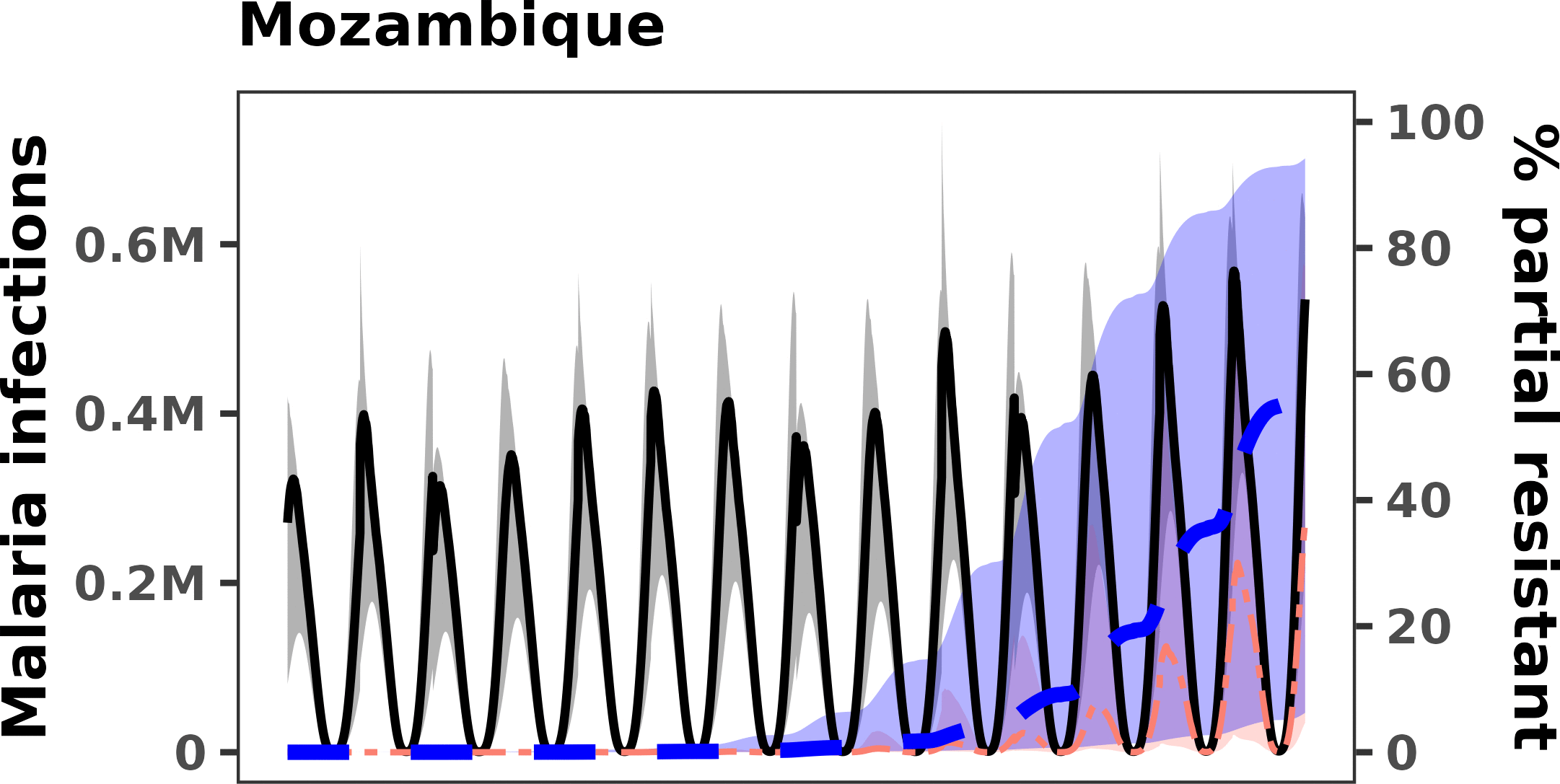

### S4 Fig 4e

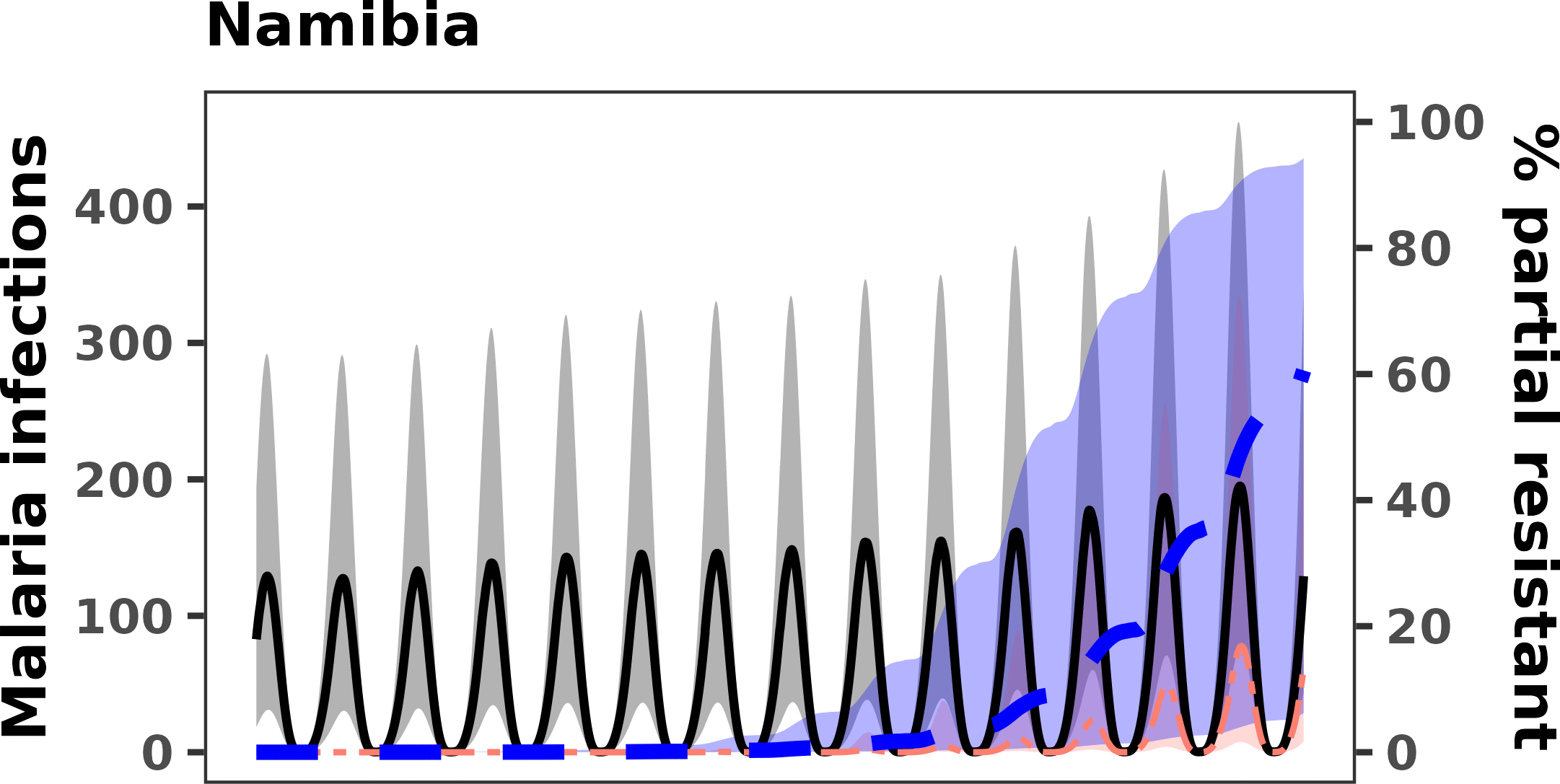

### S4 Fig 4f

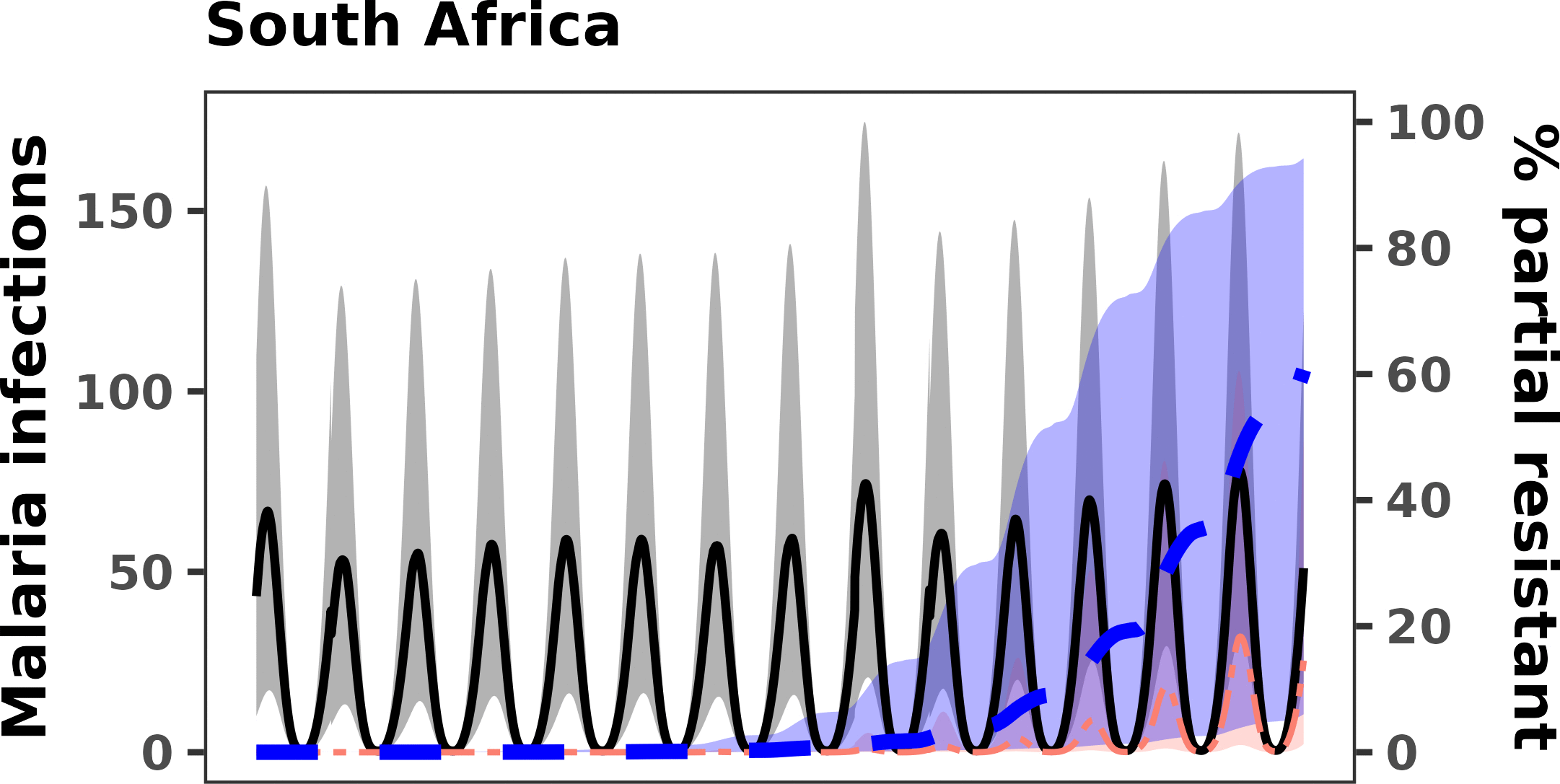

### S4 Fig 4g

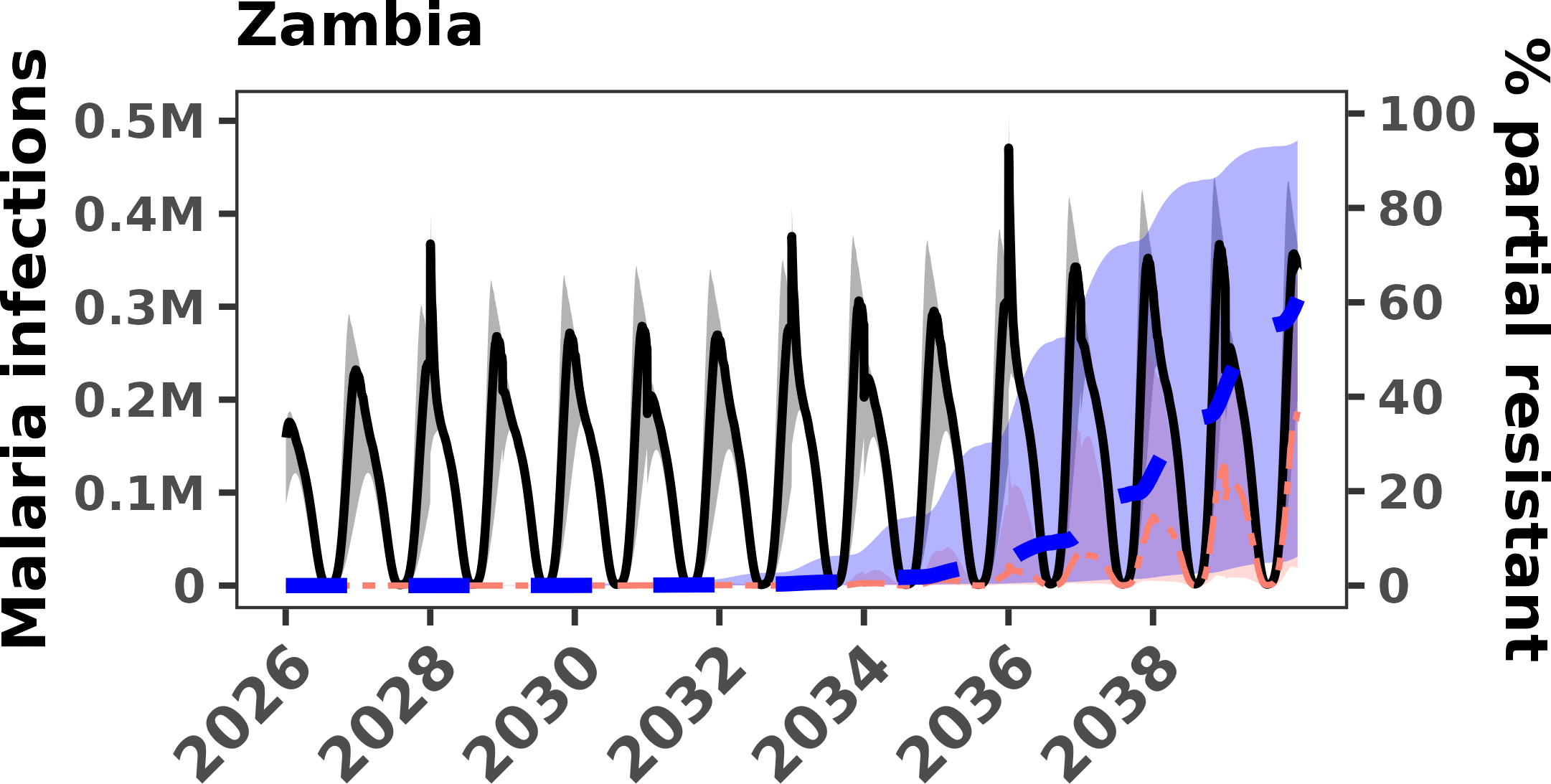

### S4 Fig 4h

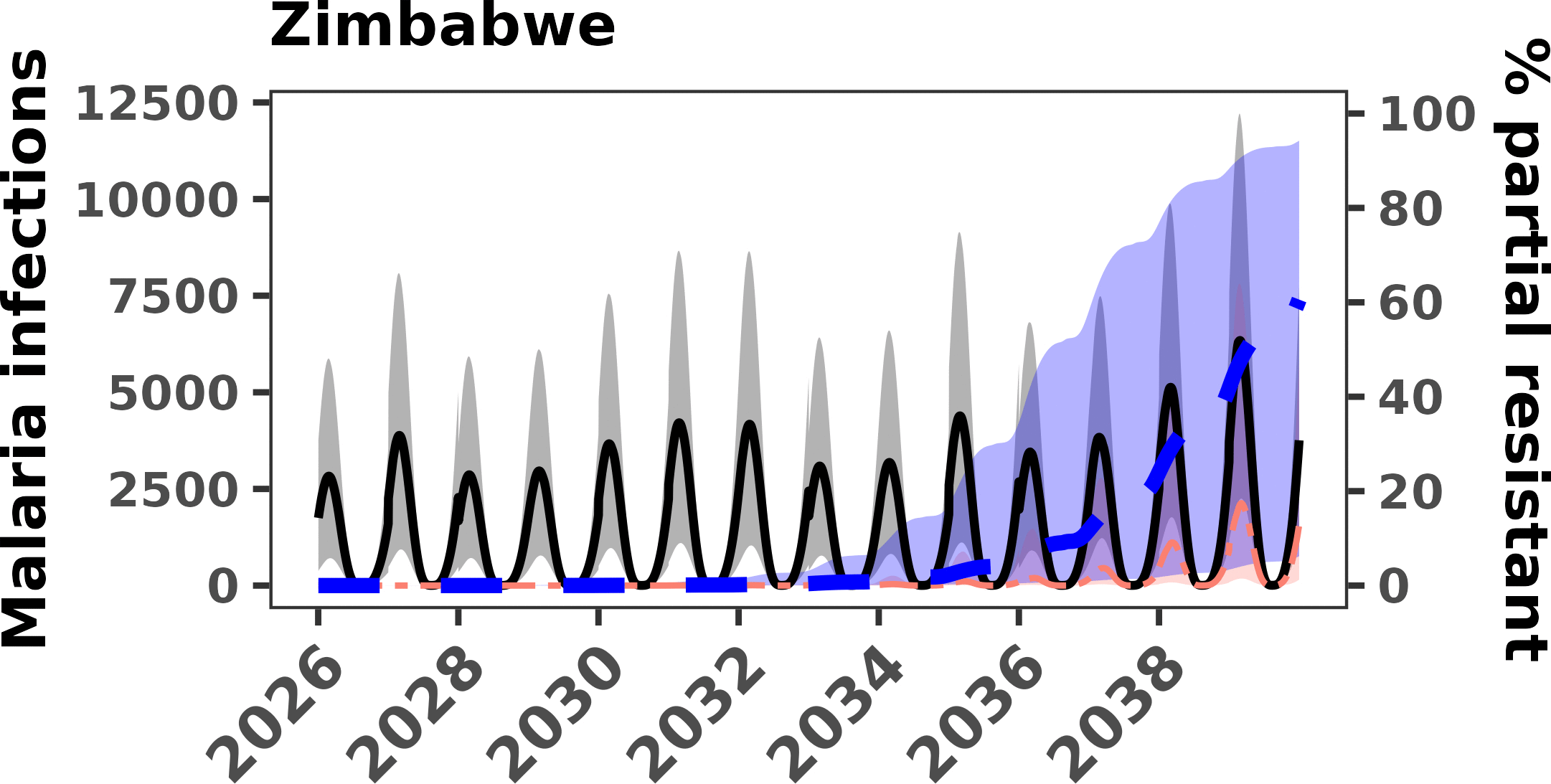

### S5 Fig 5

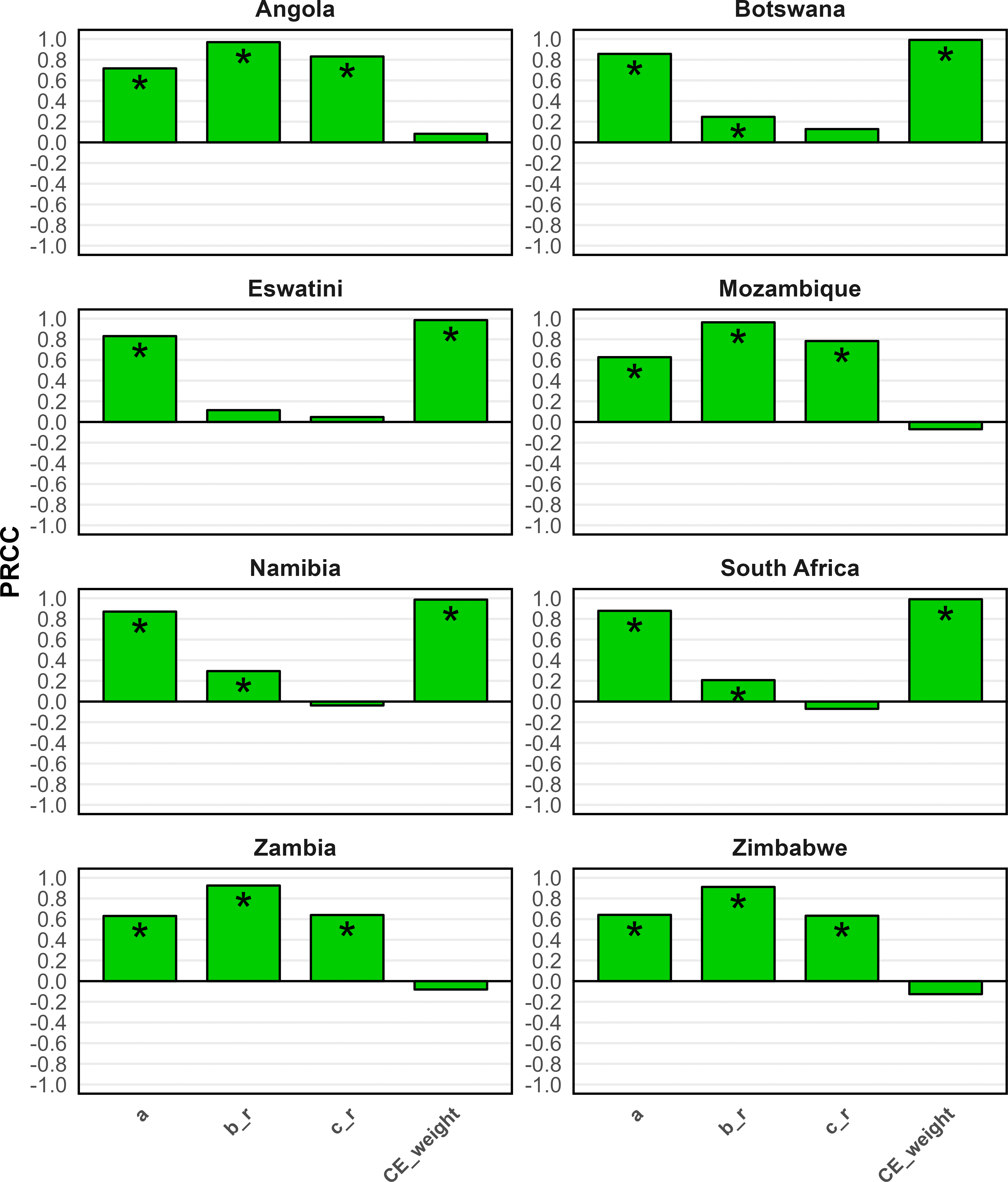
